# Navigating uncertain illness trajectories for young children with serious infectious illness: a mixed-methods modified grounded theory study

**DOI:** 10.1101/2021.07.06.21259650

**Authors:** Sarah Neill, Lucy Bray, Bernie Carter, Damian Roland, Enitan D Carrol, Natasha Bayes, Lucie Riches, Joanne Hughes, Poornima Pandey, Jennifer O’Donnell, Sue Palmer-Hill

## Abstract

Infectious illness is the biggest cause of death in children due to a physical illness, particularly in children under five years. If mortality is to be reduced for this group of children, it is important to understand factors affecting their pathways to hospital.

The aim of this study was to retrospectively identify organisational and environmental factors, and individual child, family, and professional factors affecting timing of admission to hospital for children under five years of age with a serious infectious illness (SII).

**Methods:** An explanatory modified grounded theory mixed methods design was used in collaboration with parents. Two stages of data collection were conducted: Stage 1, interviews with 22 parents whose child had recently been hospitalised with a SII and 14 health professionals (HPs) involved in their pre-admission trajectories; Stage 2, focus groups with 18 parents and 16 HPs with past experience of SII in young children. Constant comparative analysis generated the explanatory theory.

**Findings:** The core category was ‘navigating uncertain illness trajectories for young children with serious infectious illness’. Uncertainty was prevalent throughout the parents’ and HPs’ stories about their experiences of navigating social rules and overburdened health services for these children. The complexity of and lack of continuity within services, family lives, social expectations and hierarchies provided the context and conditions for children’s, often complex, illness trajectories. Parents reported powerlessness and perceived criticism leading to delayed help-seeking. Importantly, parents and professionals missed symptoms of serious illness. Risk averse services were found to refer more children to emergency departments.

**Conclusions:** Parents and professionals have difficulties recognising signs of SII in young children and can feel socially constrained from seeking help. The increased burden on services has made it more difficult for professionals to spot the seriously ill child.

## Background

Infection is a major cause of childhood deaths in the UK and globally, particularly in the under 5 year age group. The most recent analysis of child mortality data (from 2013-15) in England and Wales found that infection was associated with 20% of all childhood deaths(1). Child Death Reviews (CDR), which aim to identify modifiable factors in any child’s death, are reported by Local Safeguarding Children’s Boards and have been collated into annual reports for England by NHS Digital since 2018 and previously by the Department for Education(2). In the year ending March 2019, modifiable factors were identified in 30% of all child deaths (compared to 24% in 2016(3)) and 38% of deaths from infection(4), suggesting that more can be done to prevent these deaths.

Emergency admissions and emergency department (ED) visits have continually increased over the last 20 years. Between 1999 and 2010 emergency admissions increased particularly for under 5 year olds (<1 year by 52%, aged 1–4 years by 25%) and acute infections (by 30%)(5). This trend continued between 2007 and 2017 with a 1.6%/year increase in ED visits for all children and 3.9%/year for infants(6). In one Midlands region in the UK, 28,929 children (27.9% of all admissions) were admitted with infectious illness between 2011-2014, the largest group of emergency hospital admissions by International Classification of Diseases (ICD) coding(7). There is no single code available to indicate serious infectious illness (SII) – the focus of this paper – making it difficult to determine the exact pattern of attendance or admissions for children diagnosed with a SII.

More problematic is determining how many children’s serious illness could have been recognised sooner in primary care. These cases where the seriousness of these children’s illnesses was missed should be reported as patient safety incidents through the National Reporting and Learning System (NRLS); however, there are few reports submitted to the NRLS from primary care leading to limited learning about influences on pre-hospital care. These systems depend on recorded data; consequently, human factors are rarely captured. Notably, families’ perspectives are absent from the data collected and parents report difficulties in securing the engagement of health services in learning from their children’s deaths (www.mothersinstinct.co.uk).

The aim of our study was to retrospectively identify organizational and environmental factors and individual child, family and professional factors affecting timing of admission to hospital for children under 5 years of age with serious infectious illness (SII) in two counties in the United Kingdom. Our research questions were:

1. What, if any, social and/or personal child and family characteristics influence the journeys of children with serious infectious illness from home to hospital admission?
2. What, if any, modifiable organizational, environmental and individual human factors within health services affect the timing of the journeys of children with serious infectious illness from home to hospital admission?

### Methodological approach

Working with parents we co-designed a modified grounded theory (8, 9) explanatory, mixed-methods study (See Fig 1). Each step influenced the next and vice versa until a core category and theory which explained the findings was identified. At this stage the emerging theory was compared with existing knowledge to explore how extant evidence fitted and to identify new knowledge. This process generated our emergent theory and our findings.

**Figure 1.**
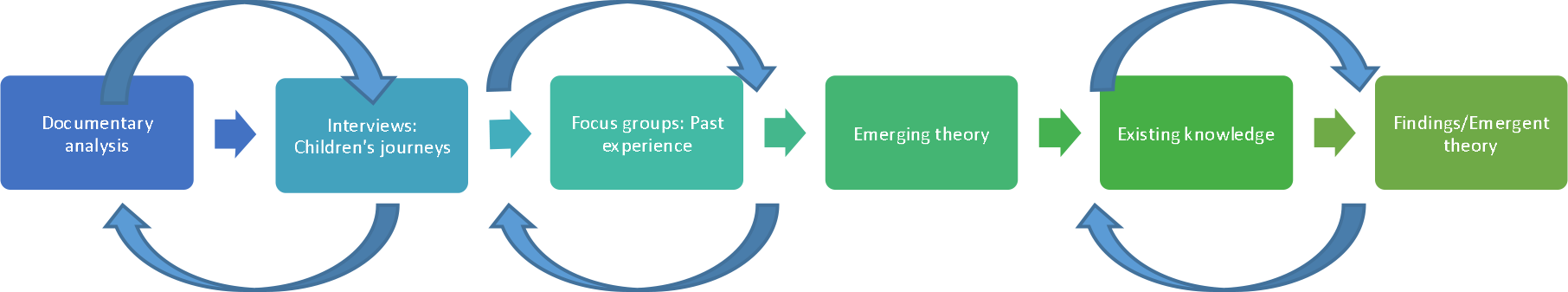
Explanatory mixed methods modified grounded theory design.

## Method

Two study areas were selected for the project representing a population served by a District General Hospital (DGH) and a Teaching (Tertiary) Hospital (TH). These two areas included patterns of service provision and population demographics similar to that in England as a whole. Ethics approval was granted by East Midlands – Nottingham 1 Research Ethics Committee (17/EM/0334) on 8 ^th^ November 2017.

Our first step was to gather available documentary evidence in each of the two study areas to provide the context for the research. The aim of this stage was to:

- identify known modifiable organizational, environmental and human factors from reports concerned with child deaths;
- gather data on patterns of service use from Hospital Episode Statistics (HES) data and ambulance service data for the preceding two years; and
- map the services available to children.

No information was available to the study team concerning learning from child death reviews in either area, consequently we were not able to analyse our data for any related information. Urgent and emergency care services were identified in each study area from health service webpages. Coding used to categorise ambulance service use for children with acute inflections was identified in collaboration with ambulance service staff so that the number of calls in each area could be identified for these children for the years 2015/16 and 2016/17. A researcher (KWD) worked with Principal Investigators (PIs) for each area to identify relevant HES coding for children presenting to hospital with a serious infectious illness so that data from the two hospitals could be compared. These codes are based on diagnostic classifications and record an episode of continuous care, consequently the data does not identify the numbers of children but does provide data on the level of activity in each hospital. Data were analysed using descriptive statistics to identify any differences between the two study areas. For further information on the documentary analysis please see S1 Fig.

Our next steps were to undertake data collection in two stages. Stage 1 involved in-depth interviews with families whose child had recently been treated for a SII in one of the two hospitals in our study area and the health professionals involved in their pre-hospital admission journeys. Stage 2 involved focus groups with parents (recruited nationally) and professionals (recruited in the area surrounding the two study sites) who had experience of child(ren) with SII between 2011 and 2018. Parents recruited to the focus groups provided data concerning their memories of these traumatic events and how these longer term memories had influenced their future health service use. HPs in Stage 2 were all in clinical practice at the time so had recent and longer term experiences to share.

These stages aimed to provide a comprehensive examination of the journey children with a SII travelled from falling ill at home to being admitted to hospital. We included families with children under 5 years of age who had had a SII, excluding neonates less than 28 days of age, post-neonatal babies who had never left hospital, children who died at home, children in receipt of palliative care or whose death was expected prior to the infection and children living outside either hospitals’ catchment areas. We were unable to identify a pre-existing definition for SII to adopt for our study.

Consequently, based on expert opinion of clinicians in the study team (DR, EC, PP), within this study we considered children to have had a serious infectious illness if they had received care on a paediatric intensive care (PICU) or high dependency unit (HDU) for a minimum of 48 hours with a diagnosis of infection. Our methods and approaches were guided by our parent collaborators.

### Recruitment

In Stage 1, families were recruited between January and 2018 and Oct 2019 and March 2019 in the hospital setting by clinical research nurses once their child was improving and had been transferred from PICU/HDU to a children’s ward: three from the DGH and nine from the TH. These families were followed up by phone at home after discharge from hospital, by member of the research team (SN, KWD). Informed consent was obtained face-to-face at the beginning of the interviews. All the family member participants were parents or primary carers of the children concerned. Throughout this paper the term parent is used to refer to all of the parent and carer participants. During the interview, parents were asked for permission to contact the health professionals involved in their child’s care. These professionals were then contacted by a researcher (KWD), given information about the project and invited to take part in the project.

Parent participants in Stage 2 were recruited through a local parent panel, by word of mouth and Facebook and through our charity partners between May and October 2019. Posters and leaflets for GP practices disseminated through primary care networks generated no interest. Health professional participants were recruited by members of the research team (DR, KWD, PP) and the local clinical research network by email and word of mouth.

### Data collection

The first stage of data collection involved retrospective in-depth interviews with parents of children under 5 years whose child had been discharged from hospital within the last 4 weeks following treatment for a serious infectious illness (SN, KWD). These audio-recorded in-depth interviews were conducted in the family home. Parents were asked to ‘*Tell me the story of your child’s illness from the time you first noticed something was wrong up until they were admitted to hospital?*’ followed by neutral prompts to help them tell us more about their experiences.

We then interviewed HPs who had been involved in these children’s pre-hospital journeys. All the HPs were interviewed by KWD in person within a quiet room in their workplace. Each HP was asked to ‘*Tell me the story of the child’s illness during the time they were in your care*’ followed by neutral prompts to generate further detail.

The second stage of data collection involved three focus groups with parents whose child had had a SII between 2011 and 2018 from across the UK in locations away from health services. A further three focus groups were held in hospital seminar rooms with HPs from the area surrounding the study sites who had experience of caring for such children in first contact services during the same time period. Each focus group was audio-recorded and facilitated by two people from the research team (KWD, SN, TB) and on one occasion a clinical research nurse from the TH. Parent focus groups were asked the starter question: ‘*Thinking back about your child’s illness, what helped or prevented you getting them admitted to hospital quickly?*’ Health professional focus groups were asked a similar starter question: ‘*What do you think are the key factors influencing the timing of admission to hospital for children with serious infectious illness?*’. These questions were followed by a series of questions that had arisen from analysis of the Stage 1 data creating a semi-structured discussion.

### Data analysis

Data were analysed inductively (no a priori coding) using the constant comparative method(10), including line by line coding facilitated through the use of QSR NVivo 11 and drawing timeline diagrams depicting each child’s pathway to hospital admission (SN, LB). Data from our documentary analysis were combined with the analysis of the interview and focus group data – in Glaserian grounded theory both qualitative and quantitative data can be used to develop theory reflecting Glaser’s mantra ‘*all is data*’ (11 p145). Glaser’s 6 Cs coding frame(8) facilitated the identification of, and interrelationships between, factors influencing children’s pathways. In common with most grounded theory research projects, we did not identify any covariances (when two variables change at the same time), making ours a 5 Cs model of Context, Conditions/Antecedents, Causes, Contingencies/Influencing variables and Consequences, all of which related to A, the Core category (Fig 2).

**Figure 2.**
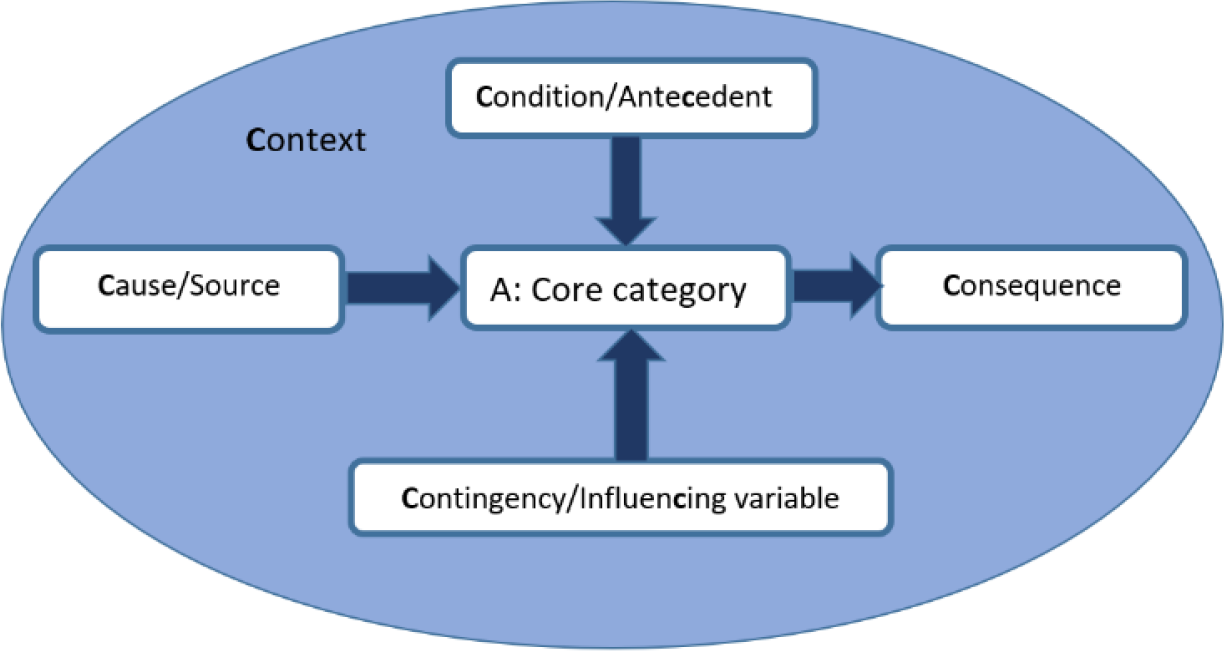
5C’s coding family adapted from Glaser’s 6Cs.

A core category is central to the data, accounting for a large proportion of the variation in behaviour as all the other categories are related to it within, what is now, the identified theory (8, 10). Once the emerging theory had been identified, its fit with existing knowledge(12), including our systematic literature review(13), was explored. Saturation was considered to have been achieved as ‘*the theory is abstract and linked to the literature, the findings are generalizable to new incidents, and the findings surprise and delight the reader*.’(14). The outcome of this final process is the theory represented in Fig 3 ‘Navigating uncertain illness trajectories: relationships between categories’.

**Figure 3.**
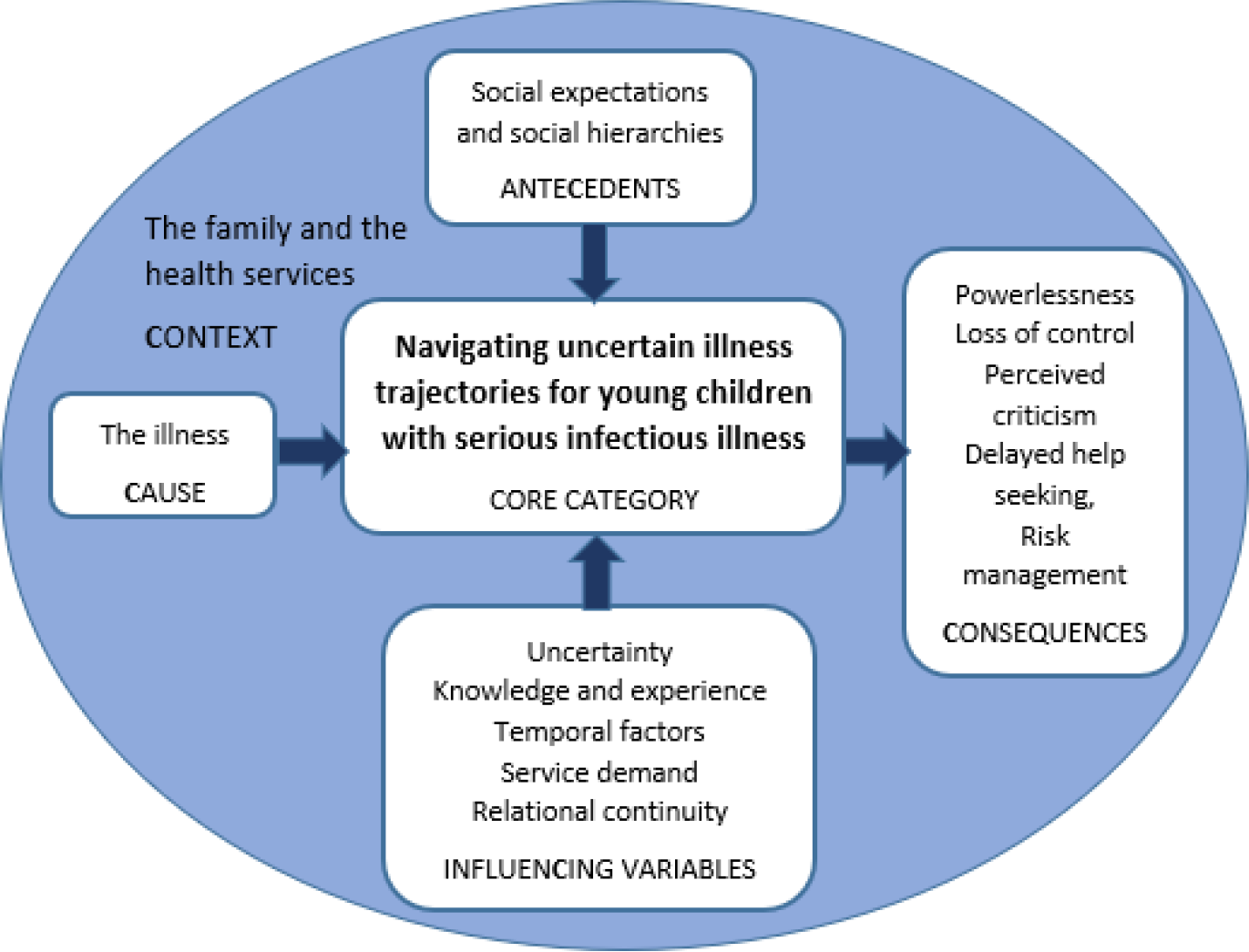
Navigating uncertain illness trajectories: relationships between categories.

## Findings

### Study participants

A total of 70 individual participants were recruited to the project between January 2018 and October 2019. In Stage 1 twelve families (a total of 22 parents), three from the DGH and nine from the TH (Table 1), and 14 health professionals (Table 2) were recruited.

**Table 1.** Stage 1 Characteristics of parent/carer participants and their affected child (N=22∼)

| Characteristic | Number of parents (%) | Characteristic | Number of parents (%) |
| --- | --- | --- | --- |
| **Age** |  | **Relationship to the child** |  |
| 25-29 years | 3 (13%) | Parent: Mother | 11 (50%) |
| 30-39 years | 10 (44%) | Parent: Father | 8 (36%) |
| 40-49 years | 0 | Other family carer | 3 (14%) |
| 50-59 years | 1 (4%) |  |  |
| 60+ years | 3 (13%) |  |  |
| **Gender** |  | **Income** |  |
| Female | 12 (52%) | Less than 10,000 | 3 (13%) |
| Male | 9 (39%) | 10,000-19,999 | 5 (22%) |
| **Ethnicity** |  | 20,000-29,999 | 4 (17%) |
| White British | 12 (52%) | 30,000-39,999 | 5 (22%) |
| Indian | 6 (26%) | 40,000-49,999 | 0 |
| **Employment** |  | 50,000-59,999 | 2 (9%) |
| Employed (part or full time) | 8 (35%) | 60,000-79,999 | 2 (9%) |
| Unemployed or retired | 3 (13%) | 80,000-99,999 | 1 (4%) |
| Caring for family at home | 5 (22%) | 100,000+ | 3 (13%) |
| **Age of affected child\*** |  | **Diagnoses of affected child\*&\*\*** |  |
| Under 6 months | 1 (8%) | Acute Respiratory | 12 (52%) |
| 6-12 months | 2 (17%) | Acute exacerbation of recurrent respiratory | 5 (22%) |
| 13-23 months | 2 (17%) | Acute disseminated encephalomyelitis (ADEM) | 1 (4%) |
| 2-4 years old | 7 (58%) | Tonsillitis | 1 (4%) |
|  |  | Sepsis and Septicaemia | 2 (9%) |
| ~Although 22 parents/carers completed the questionnaire, questions were not compulsory and therefore each question was not always completed by 100% of parents. |  |  |  |
| *Based on the number of families (N=12) engaged in Stage 1, not on the total number of parents (N=22) participating in Stage 1. |  |  |  |
| **Many children had multiple diagnoses. |  |  |  |

**Table 2.**
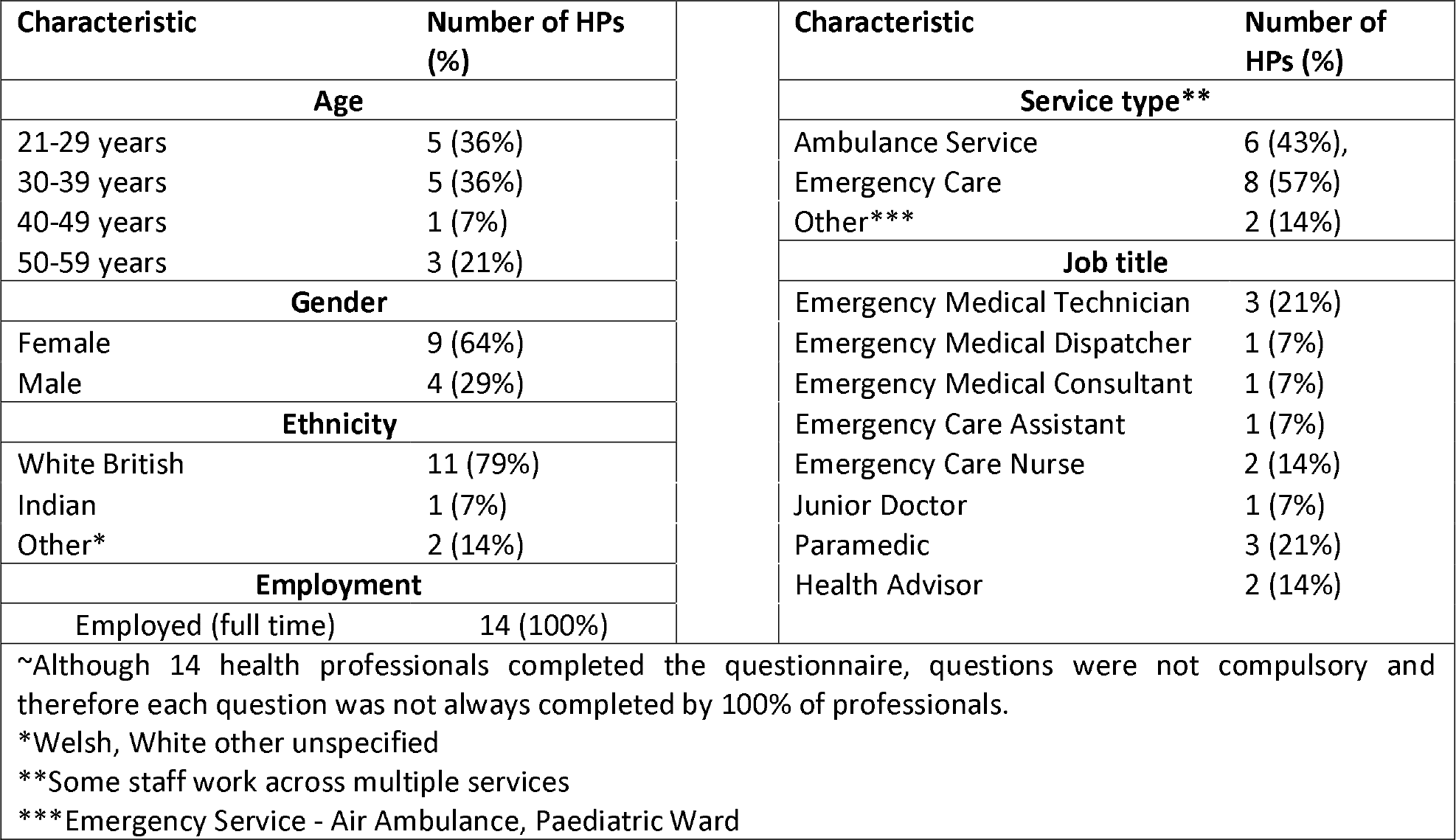
Stage 1 Characteristics of Health professional (HP) participants (N=14∼)

In Stage 2, a total of 18 parents (Table 3) and 16 HPs (Table 4) were recruited. Health professionals were from our study area, but as local recruitment of parents generated only two participants, we recruited nationally through our charity partners for the parent focus groups. Six parents were unable to attend the focus groups, opting to take part in individual telephone or email interviews.

**Table 3.** Stage 2 Characteristics of parent participants (N=18∼)

| Characteristic | Number of parents (%) | Characteristic | Number of parents (%) |
| --- | --- | --- | --- |
| **Age** |  | **Relationship to the child** |  |
| 30-39 years | 11 (61%) | Parent: Mother | 15 (83%), |
| 40-49 years | 5 (28%) | Parent: Father | 2 (11%) |
| **Gender** |  | **Income** |  |
| Female | 14 (78%) | Less than 10,000 | 2 (11%) |
| Male | 2 (11%) | 10,000-19,999 | 3 (17%) |
| **Ethnicity** |  | 20,000-29,999 | 0 |
| White British | 12 (67%) | 30,000-39,999 | 0 |
| White other* | 3 (17%) | 40,000-49,999 | 1 (6%) |
| **Employment Status** |  | 50,000-59,999 | 1 (6%) |
| Employed (part or full time) | 12 (67%) | 60,000-79,999 | 3 (17%) |
| Unemployed | 1 (6%) | 80,000-99,999 | 4 (22%) |
| Caring for family at home | 3 (17%) | 100,000+ | 2 (17%) |
| **Age of affected child\*\*** |  | **Diagnoses of affected child\*\*&\*\*\*** |  |
| Under 6 months | 6 (38%) | Acute Respiratory | 1 (6%) |
| 6-12 months | 4 (25%) | Sepsis and Septicaemia | 6 (38%) |
| 13-23 months | 2 (12%) | Meningitis | 14 (88%) |
| 2-4 years old | 4 (25%) |  |  |

~Although 18 parents completed the questionnaire, questions were not compulsory and therefore each question was not always completed by 100% of parents. \*European, Scottish, Other unspecified. \*\*Based on the number of families (N=16) engaged in Stage 2, not on the total number of parents (N=18) engaged in Stage 2. \*\*\*Many children have multiple diagnoses.

**Table 4.** Stage 2 Characteristics of health professional (HP) participants (N=16∼)

| Characteristic | Number of HPs (%) | Characteristic | Number of HPs (%) |
| --- | --- | --- | --- |
| **Age** |  | **Service type** |  |
| 21-29 years | 2 (13%) | General Practice | 5 (32%) |
| 30-39 years | 6 (38%) | Emergency Care | 5 (32%) |
| 40-49 years | 4 (25%) | Ambulance Service | 2 (13%) |
| 50-59 years | 4 (25%) | Other** | 4 (25%) |
| **Gender** |  | **Job title** |  |
| Female | 9 (56%) | General Practitioner | 5 (32%) |
| Male | 5 (32%) | Paediatric Emergency Medical Consultant | 4 (25%) |
| **Ethnicity** |  | Emergency Care Children's Nurse | 1 (6%) |
| White British | 10 (63%) | Community Children's Nurse | 1 (6%) |
| South Asian* | 3 (19%) | Paramedic | 2 (13%) |
| African | 1 (6%) | Other*** | 3 (19%) |
| Other* | 2 (13%) |  |  |
| **Employment** |  |  |  |
| Employed (full time) | 12 (75%) |  |  |
| Employed (part time) | 4 (25%) |  |  |

~Although 16 health professionals completed the questionnaire, questions were not compulsory and therefore each question was not always completed by 100% of professionals. \*Indian, Pakistani, Bangladeshi \*\* NHS111, Community \*\*\*Community Pharmacist, Dental Hygienist Oral Health Lead, Health Advisor

### Navigating uncertain illness trajectories for young children with serious infectious illness: The emergent theory

From the onset of the illness, uncertainty ran throughout parents’ and health care professionals’ stories of navigating social expectations and hierarchies and health services to enable these children to access appropriate treatment in a timely manner. Parents reported trying to navigate multiple pathways though complex services whilst also having to overcome perceived criticism of their behaviour and decision making. Heath care professionals also reported the need to navigate complex health services and social hierarchies between professional groups. This uncertainty in many cases delayed help seeking or referral. If the NHS is conceptualised as a safety net designed to promote health and prevent avoidable morbidity and mortality, most of the children in this study have fallen, at least in part, through this safety net.

The interrelated sub-categories that make up the emergent theory are presented below with a ‘C’ used to highlight which of Glaser’s 6 Cs these represent. Categories are presented beginning with ‘The Illness’, the Cause category in grounded theory terms, followed by ‘Navigating uncertain illness trajectories’, the Core category to which all the other categories relate, then ‘The family and the health services Context’ within which these trajectories took place, the ‘Social expectations and social hierarchies’, the anteCedents or Conditions, the ‘InfluenCing variables or Contingencies’ affecting these trajectories and finally the ‘Consequences’ of these complex illness trajectories.

Throughout the presentation of the findings, participants are referred to using unique codes (see Box 1).

#### Box 1 Participant codes

| Research stage | Type of participant | Code | Additional information |
| --- | --- | --- | --- |
| Stage 1 | Parents | Study site followed by P for parent and family number (e.g. TH,P01). | Teaching Hospital = TH; District General Hospital = DGH |
|  | Health professionals | HP followed by a number and service identifier such as NHS111/GP/Amb.tech/ED nurse/999 call handler (e.g. HP01, NHS111). | NHS 111 (NHS24 in Scotland) is a non-emergency medical helpline free to use in the UK.<br>GP is the accepted abbreviation for general practitioners – family doctors in the UK.<br>Amb.tech is short for ambulance technician – they work with paramedics on ambulances but have less training.<br>999 is the telephone number for the UK’s emergency service. |
| Stage 2 | Parents | P for Parents, FG for focus group, FG number (e.g. FG1), M for Mother or F for Father followed by respondent number (e.g. P,FG2,M7).<br>Parents who contributed by telephone are identified as P,FGT.<br>Parents who contributed by email as P,FGE (e.g. P,FGT,M1 or P,FGE,M2). | Three parent focus groups<br>P,FG1<br>P,FG2<br>P,FG3 |
|  | Health professionals | Location of the focus group i.e. TH or DGH followed by HP,FG and the number of the focus group (e.g. TH,HP,FG2). | Individuals are identified by appending their service identified as for Stage 1. |

### The illness: the Cause and beginning of the illness trajectory

The beginning of all the children’s journeys was the onset of illness. Of the 28 children whose parents shared illness trajectories with researchers, 10 children from Stage 1 were reported to have a respiratory illness, one had tonsillitis and one had acute disseminated encephalomyelitis (ADEM) (see Table 5).

**Table 5.** Stage 1 Characteristics of each family and affected child. TH Teaching hospital; DGH District general hospital; NP Nurse Practitioner; CAU Child Assessment Unit.

| Stage 1 Case | Family members interviewed | Age band of affected child | Household composition | Pre-existing conditions (yes/no) | Diagnosis for this illness | Duration of this illness prior to admission | Services accessed pre-hospital and admitting unit |
| --- | --- | --- | --- | --- | --- | --- | --- |
| THP04 | Mother | 13-23 months | Two parents | Yes | ?Bronchiolitis | 3 + days | GP, CAU, Ambulance, ED, HDU |
| THP05 | Father | Under 6 months | Two parents; 6 other adults and their 4 children | Unknown | RSV Bronchiolitis and Influenza A | Approx. 7 days | GP x3, EDx2, CAU, PICU |
| THP08 | Mother and Father | 2-4 year old | Two parents, grandparent and one young sibling. | Yes | ?Chest infection | Approx. 6 days | GP, Ambulance, ED, PICU |
| THP10 | Mother and Father | 2-4 year old | Two parents and one younger sibling. | Yes | ?Asthma attack and chest infection | 1.5 days | NP at GP surgery, Ambulance, ED, PICU |
| THP12 | Mother | 2-4 year old | Two parents and two older siblings. | Yes | Asthma attack and chest infection | Approx. 12 hours | NP at GP surgery x2, ED, HDU |
| THP18 | Mother, two other family carers | 2-4 year old | Two parents, one younger and one older sibling. | No | *'Chest infection and later pneumonia, fluid around the lung and Strep A blood infection'* | 2.5 days | NHS 111, Ambulance, ED, HDU/PICU |
| THP21 | Mother and Father | 2-4 year old | Two parents and two older siblings. | Unknown | ADEM - Acute disseminated encephalomyelitis | 6 days | GP x2, ED x2, Walk-in Centre, ED, HDU/PICU |
| THP22 | Mother and Father | 2-4 year old | Two parents | Yes | Tonsillitis with obstruction | 7 days | Walk-in Centre, locum GP, NHS 111, Ambulance, ED, PICU |
| THP27 | Mother and one other family carer | 6-12 months | Two parents and one older sibling. | Yes | Bronchiolitis (recurrence) with obstruction | 10 days | Ambulance, ED, PICU |
| DGHP01 | Mother and Father | 13-23 months | Two parents. | No | Collapsed lung and sepsis | 12 days | GP x3, NHS 111, ED, HDU |
| DGHP02 | Mother and Father | 6-12 months | Two parents and one older sibling. | No | Collapsed lung secondary to ?chest infection/ pneumonia | Approx. 8 days | GP x2, NHS 111, Ambulance, ED, HDU |
| DGHP03 | Mother and Father | 2-4 year old | Two parents. | No | Pneumonia | 7 days | GP, NHS 111, Ambulance, 999, ED, HDU/PICU |

In Stage 2, 14 children were reported to have meningitis (five also had sepsis), one had urinary sepsis and one had bronchiolitis (see Table 6). The high number of children with meningitis in Stage 2 reflects the success of recruitment through our charity partner, Meningitis Now.

**Table 6.** Stage 2 Characteristics of each family and affected child. P,FG1 = Parent Focus group 1, August 2019; P,FG2 = Parent Focus group 2, October 2019; P,FG3 = Parent Focus group 3, October 2019; P,FGT = Parent Focus group alternative telephone interview: October 2019; P,FGE = Parent focus group alternative email interview: October 2019; M = Mother F = Father followed by the number of the participant e.g. M1

| Stage 2 Case | Family members interviewed | Age band of affected child | Household composition | Pre-existing conditions (yes/no) | Diagnosis for this illness | Sequelae of the illness | Duration of this illness prior to admission | Services accessed pre-hospital and admitting unit |
| --- | --- | --- | --- | --- | --- | --- | --- | --- |
| P,FG1,M1 | Mother | 6-12 months | Two parents and two children. | Yes | Bronchiolitis | Unknown | Unknown | ED, HDU |
| P,FG1,M2 | Mother | 6-12 months | One parent and four children. | No | Meningitis and sepsis | Right below elbow amputee. Acquired brain injury. Stomach damage causing food sensitivities. Growth plate damage | 4 days | NHS111, Ambulance, ED, Ward |
| P,FG2,M1 | Mother | 2-4 year old | Two parents and six children. | Unknown | Meningitis | No bone growth in both legs due to sepsis. Now having treatment (lengthening and correcting the shape of the legs) | 3 days | GP, 999, ambulance, 'Hospital' |
| P,FG2,M2 | Mother | 6-12 months | Two parents and three children. | Unknown | Meningococcal septicaemia | Unknown | 24 hours | GP, ED, PICU |
| P,FG2,M3&F4 | Mother and Father | Under 6 months | Two parents and two children. | Unknown | Late onset group B streptococcus meningitis | Child died | 24 hours | GP, 999, ED, 'Hospital' |
| P,FG2,M5&F6 | Mother and Father | 2-4 year old | Two parents and two children. | Unknown | Meningitis B | Child died | < 24 hours | 999, ED, PICU |
| P,FG2,M7 | Mother | 2-4 year old | Two parents and one child. | Unknown | Meningitis | Child died | 3 days | ED, 'Hospital' |
| P,FG3,M1 | Mother | 2-4 year old | Two parents and two children. | Unknown | Meningococcal disease | Unknown | 24 hours | GP, NHS111, Ambulance, ED, PICU |
| P,FG3,M2 | Mother | 6-12 months |  | Yes | Pneumococcal meningitis | Child died | 2 weeks + | GPx4, ED, Adult HDU |
| P,FG3,M3 | Mother | 6-12 months | Two parents and two children. | Unknown | Pneumococcal meningitis | Unknown | 2 weeks + | Walk-in centre, GP, ED, 'Hospital' |
| P,FGT,M1 | Mother | 13-23 months |  | Unknown | Bacterial meningitis and septicaemia | Unknown | 2 days | OOHS GP, EDx2, 'Hospital' |
| P,FGT,M2 | Mother | Under 6 months | Two parents and two children. | Unknown | Viral meningitis | Unknown | 12 hours | NHS24, OOHS Nurse, Ambulance, ED, 'Hospital' |
| P,FGT,M3 | Mother | Under 6 months | Two parents and one child. | Unknown | Meningitis | Unknown | 12 hours | GP, ED, 'Hospital' |
| P,FGT,M4 | Mother | Under 6 months | Three adults and one child | Unknown | Meningitis and sepsis | Unknown | <24 hours | NHS111, Urgent Care Centre, 'Hospital' |
| P,FGE,M1 | Mother | Under 6 months | Two adults and four children. | Unknown | Urinary sepsis | Unknown | 6 days | HV, NHS24 x2, OOHS GP, GP, ED, 'Hospital' |
| P,FGE,M2 | Mother | Under 6 months | Two adults and three children. | Unknown | Meningitis and septicaemia | Growth plates affected result in leg length discrepancy | <24 hours | GP, GP OOHS, Cottage Hospital, Ambulance, PICU |

N.B. 'Hospital' is given as the admitting unit where no information was provided about the unit to which the child was admitted.

The duration of the illness prior to admission to hospital varied from 12 hours to 12 days in Stage 1 and from 12 hours to more than 2 weeks in stage 2 illustrating the individual and unpredictable trajectory of each child’s illness.

### Navigating uncertain illness trajectories

#### Defining the illness and its severity during the illness trajectory

Throughout the illness trajectory, parents had to make sense of the illness and its severity. Parents’ ability to define the illness and judge its seriousness was affected by tiredness, distractions of family life, past experience, knowledge of symptoms/illness and not wanting it to be serious as the ‘*thought of it being something more is unbearable*’ (P,FG2,M5). In the later stages of the trajectory towards hospital admission, parents perceived that the illness had progressed from minor to very obviously real and serious, often reported in this study as recognising *significant* differences from normal or that something was obviously ‘*not right…… he didn*’t look right (DGH,P01,M); ‘*she’s not right*’ (P,FG2,M3). Before this point lay uncertainty about the legitimacy of seeking help; it is in this uncertain part of the illness trajectory that there are opportunities for parents to access earlier treatment. For some children whose illness progresses rapidly this window of time is very short.

Some symptoms of serious illness were not recognised by parents and, in a few instances, by health professionals (Box 2). The significance of wording and phrases used by parents to describe what was worrying them about their unwell child, such as ‘*not quite herself*’ (P,FG2,M3) and ‘*not there behind the eyes*’ (P,FG2,M7), were reported by some parents to be missed by HPs. The lack of recognition of these phrases illustrates the difficulties parents had in communicating their concerns about their child’s illness in terms of symptoms that were recognised by HPs. For example, one mother explained:

“*That’s where I struggle I think, to be able to explain why I know he’s not right, but I get that a lot. I think I seem to just - it’s just in me and I can’t explain it. …. The amount of times I’ve said to him [Father], ‘He’s just not right, something’s not right but I don’t know what it is*” (DGH,P01,M).

##### Box 2: Missed symptoms of serious illness

**Box 2 Missed symptoms of serious illness**
| Symptoms not recognised by parents | Symptoms not recognised by health professionals |
| --- | --- |
| ☐ 'Bruising', 'love bite', purple mark<br>☐ Temp over 38°C in young baby<br>☐ Lack of urine<br>☐ Grunting<br>☐ Head/back pain<br>☐ Mottled skin<br>☐ Sucking in under the ribs<br>☐ Fast breathing<br>☐ Funny cry<br>☐ Staring<br>☐ Stiffness<br>☐ Non-response to paracetamol | ☐ Purple mark (NHS 24 call handler)<br>☐ Temp over 38°C in young baby (Out-of-hours service (OOHS GP))<br>☐ Lack of urine (OOHS GP)<br>☐ Grunting (ED doctor) |

#### Parent help seeking during the illness trajectory

Parents made between one and six contacts with health services during their child’s illness trajectory (see Tables 7-10). Use of the out of hours service (OOHS) was rarely reported. Various factors were reported by parents to affect children’s trajectories: access to GP appointments – “*it’s quite hard to get an appointment*” (DGH,P02,F), transport – “*We’re stuck, especially with no car*” (TH,P10,F) and proximity to services –“*it is not far. That*’s why I chose it [Urgent Care Centre]” (TH,P21,F). Psychosocial factors affecting parents’ decision making about seeking help for their child are explored in Influencing Variables below.

**Table 7.**
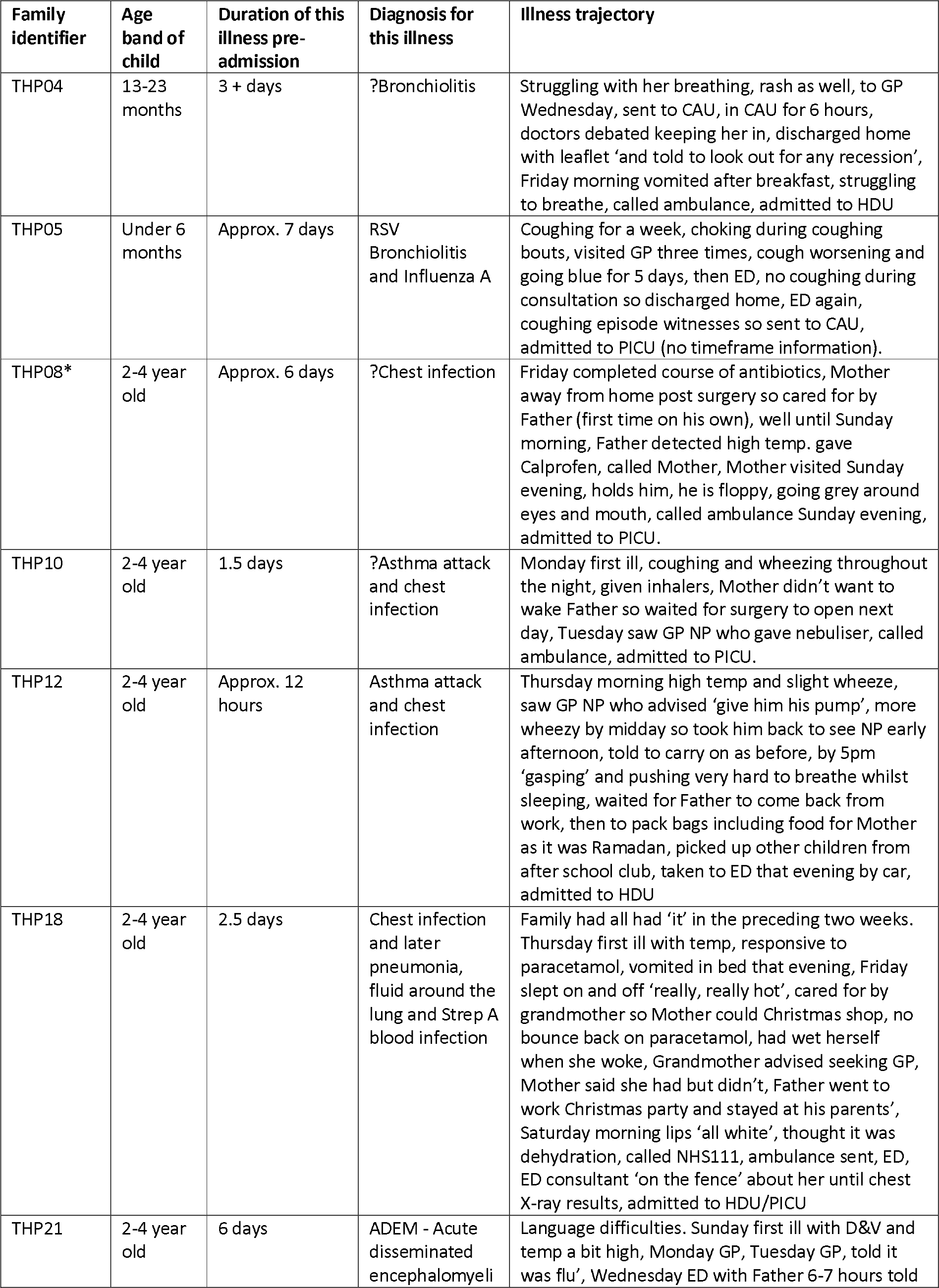
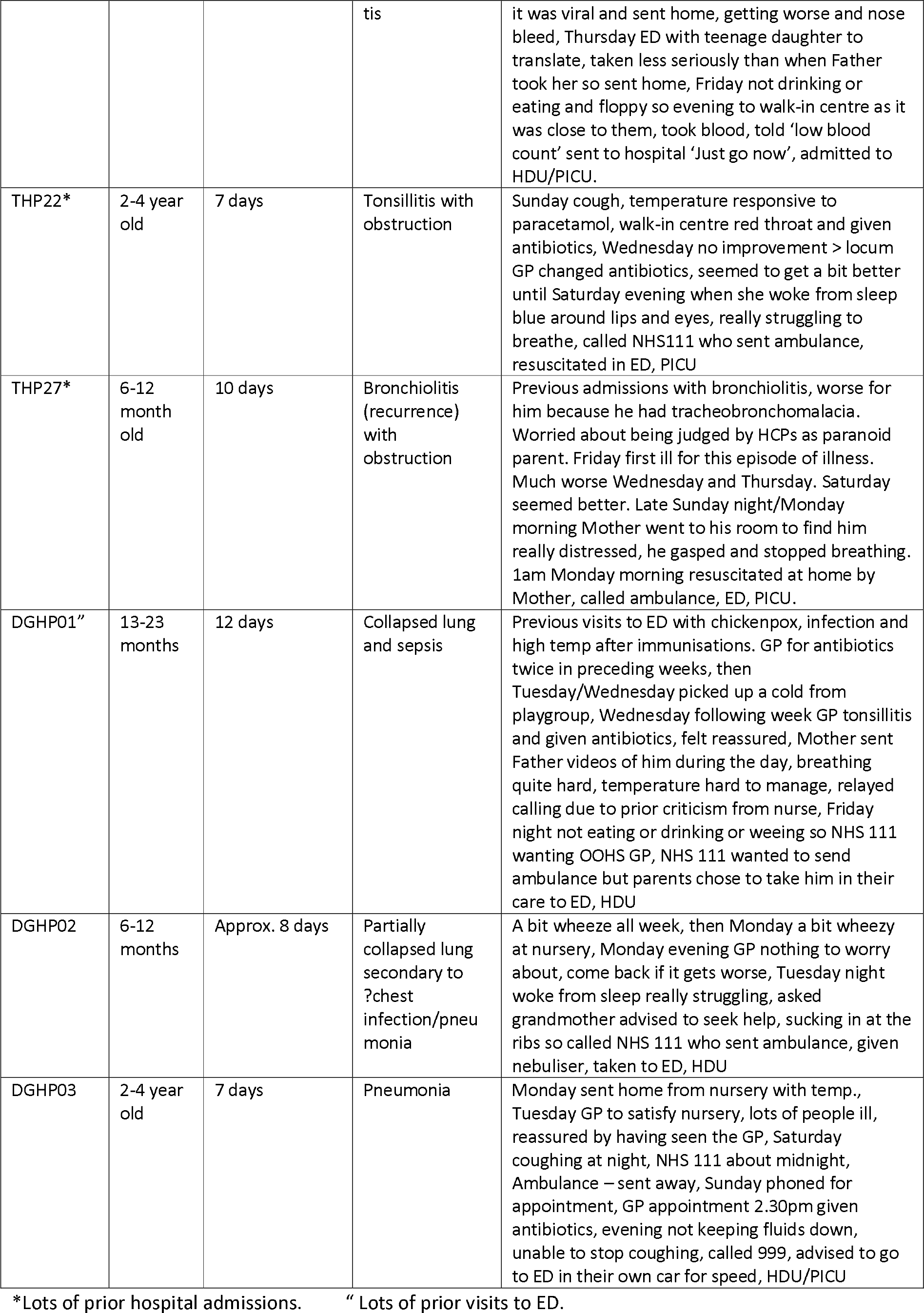
Stage1 Illness trajectories. TH Teaching hospital, DGH District general hospital, CAU Child Assessment Unit, NP Nurse Practitioner

**Table 8.**
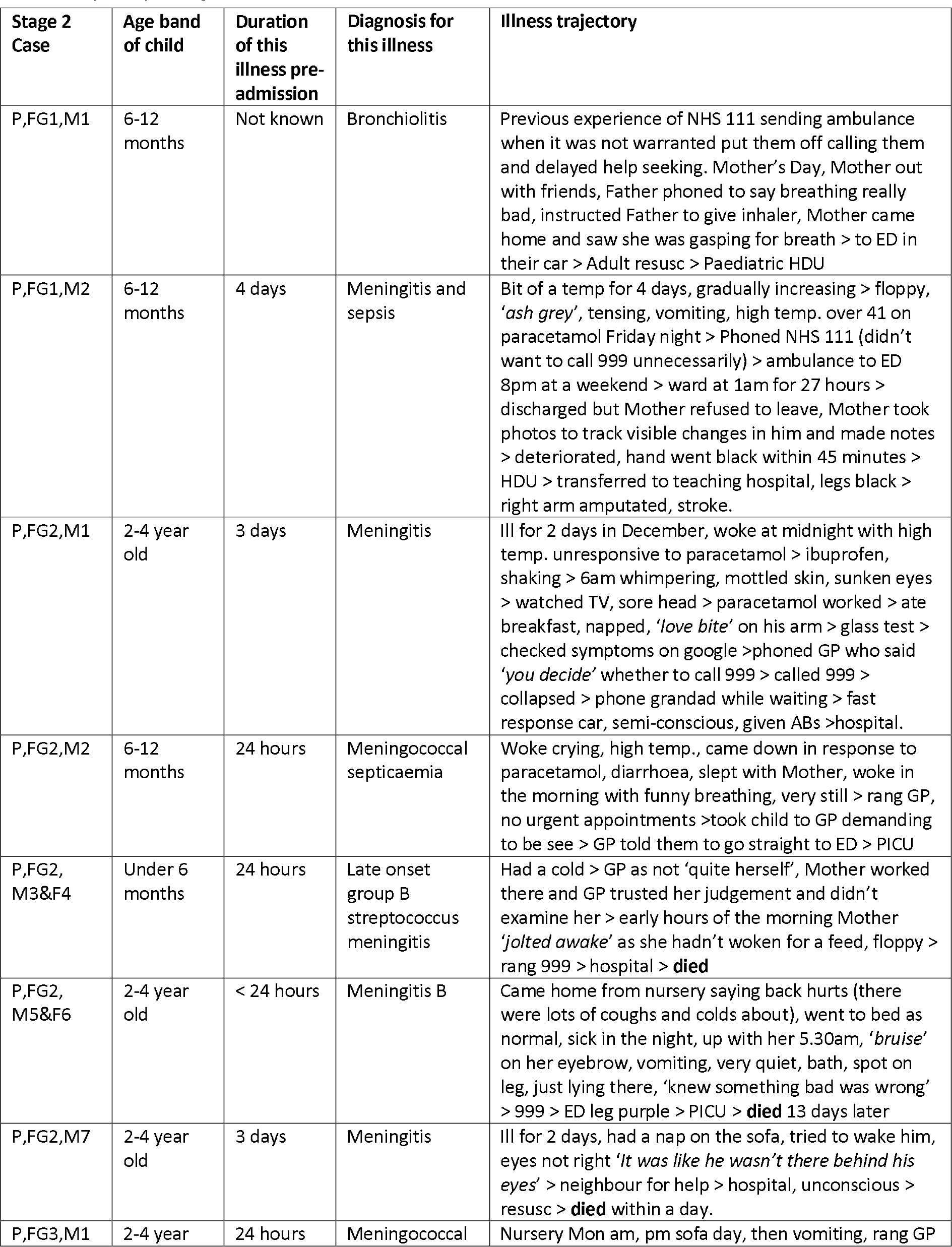
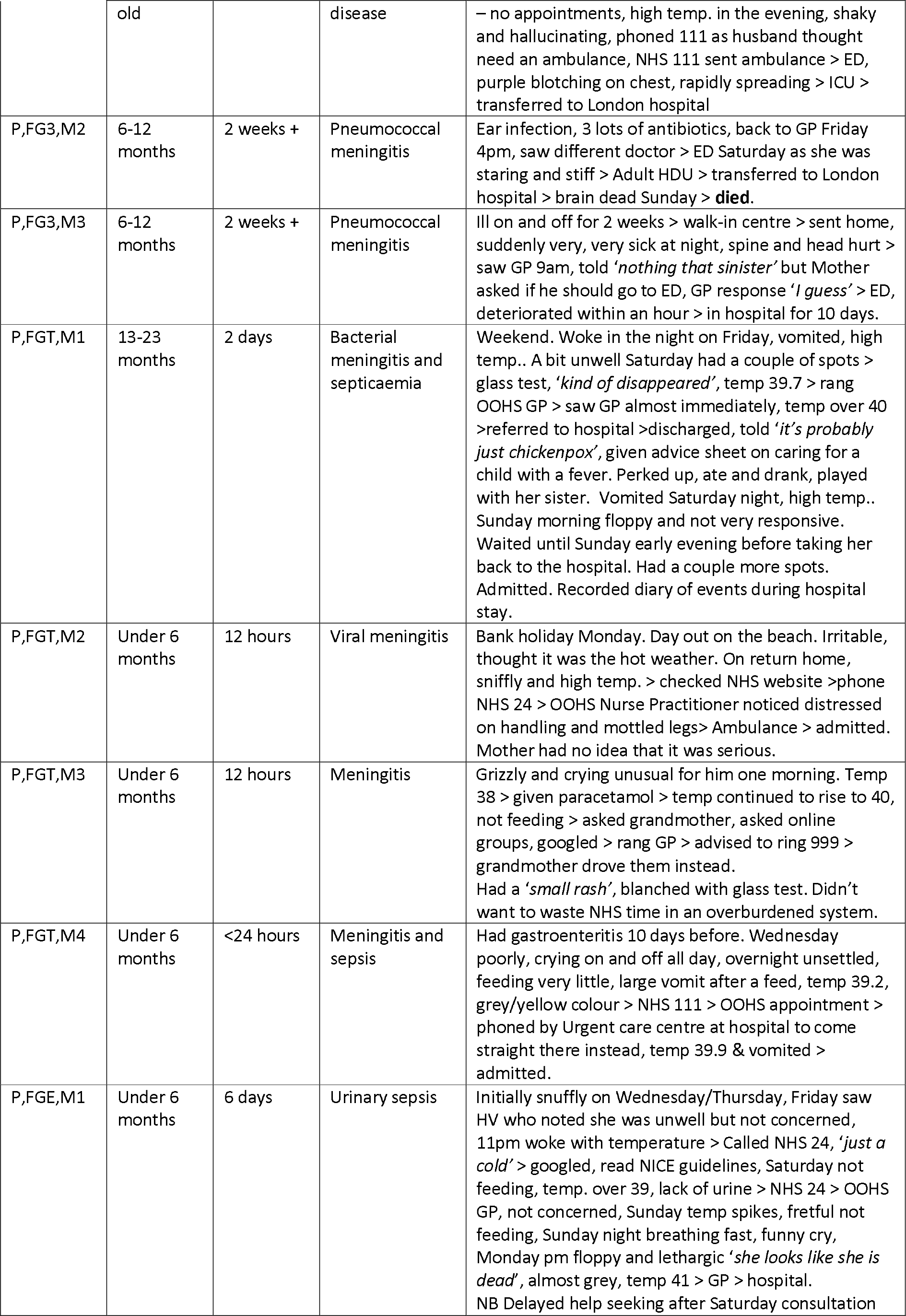
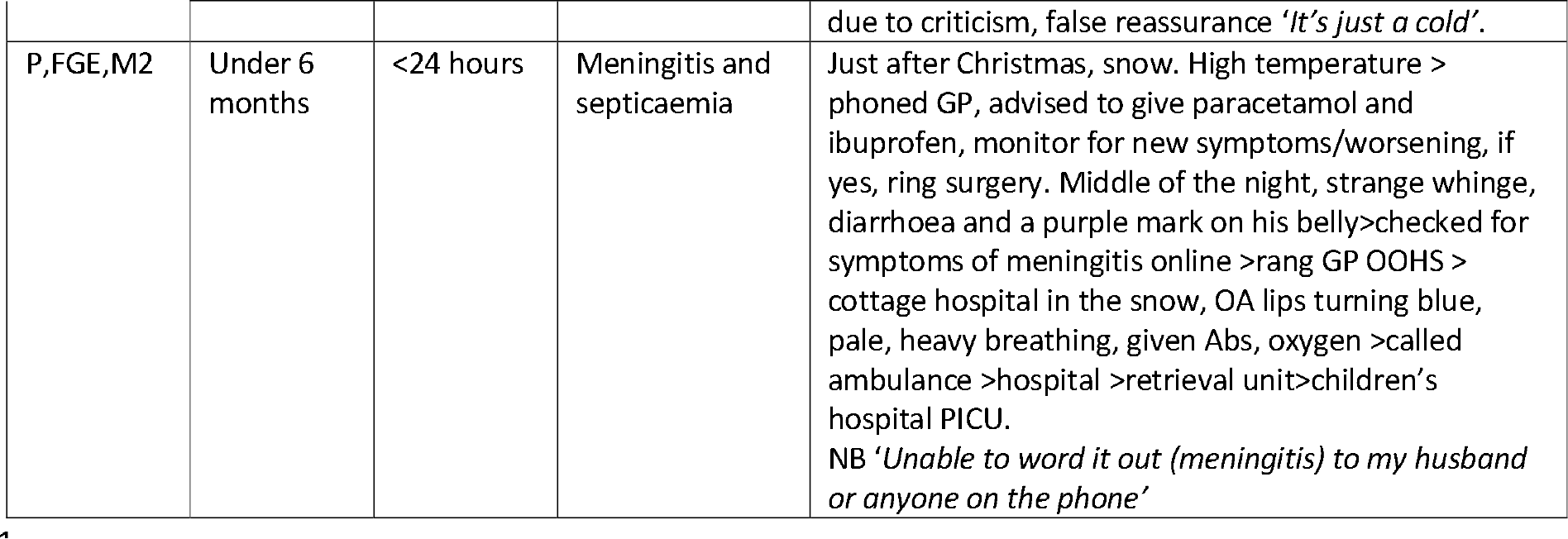
Stage 2 Illness trajectories. P,FG2 = Parent Focus group 1, August 2019; P,FG2 = Parent Focus group 2, October 2019; P,FG3 = Parent Focus group 3, October 2019; P,FGT = Parent Focus group alternative telephone interview, October 2019; P,FGE = Parent focus group alternative email interview, October 2019; M = Mother F = Father followed by the number of the participant e.g. M1

**Table 9.**
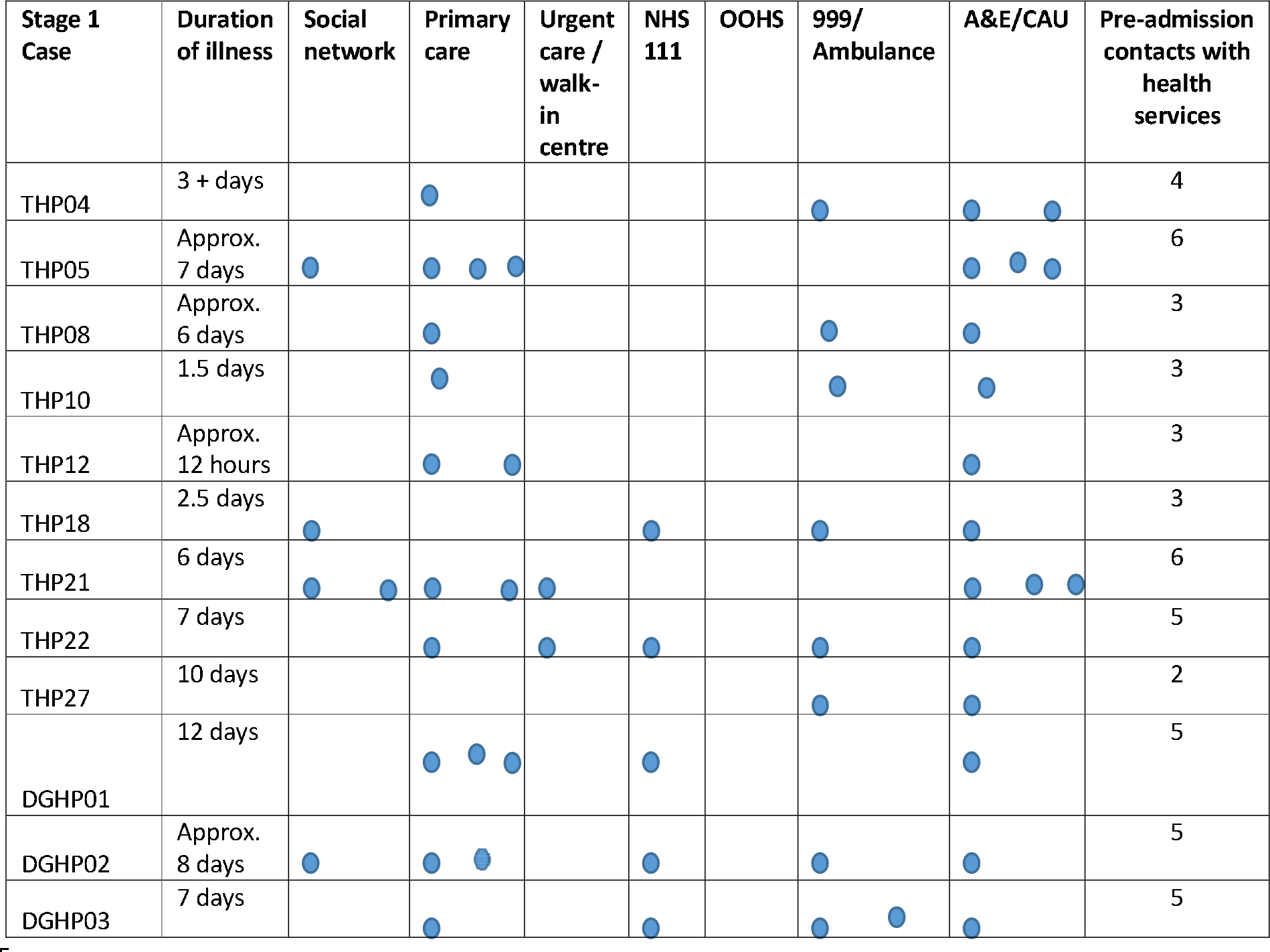
Stage 1 Children’s help seeking on their illness trajectory to hospital admission. Please note that these are not presented in the order in which parents made contact with these services THP = parent recruited in the Teaching Hospital; DGHP = parent recruited in the District General Hospital

**Table 10.** Stage 2 Children’s help seeking on their illness trajectory to hospital admission. Please note that these are not presented in the order in which parents made contact with these services. P,FG2 = Parent Focus group 1, August 2019; P,FG2 = Parent Focus group 2, October 2019; P,FG3 = Parent Focus group 3, October 2019; P,FGT = Parent Focus group alternative telephone interview, October 2019; P,FGE = Parent focus group alternative email interview, October 2019; M = Mother F = Father followed by the number of the participant e.g. M1

| Stage 2 Case | Duration of illness | Social network | Primary care | Urgent care / walk-in centre | NHS 111/ NHS24 | OOHS | 999/ Ambulance | A&E/CAU | Pre-admission contacts with health services |
| --- | --- | --- | --- | --- | --- | --- | --- | --- | --- |
| P,FG1,M1 | Not in the data | ● |  |  |  |  |  | ● | 1 |
| P,FG1,M2 | 4 days |  |  |  | ● |  |  | ● | 3 |
| P,FG2,M1 | 3 days |  | ● |  |  |  | ● | ● | 3 |
| P,FG2,M2 | 24 hours |  | ● |  |  |  |  | ● | 2 |
| P,FG2,M3&F4 | 24 hours |  | ● |  |  |  | ● | ● | 3 |
| P,FG2,M5&F6 | < 24 hours |  |  |  |  |  | ● | ● | 2 |
| P,FG2,M7 | 3 days | ● |  |  |  |  |  | ● | 1 |
| P,FG3,M1 | 24 hours |  |  |  | ● |  | ● | ● | 3 |
| P,FG3,M2 | 2 weeks + |  | ● |  |  |  |  | ● | 2 |
| P,FG3,M3 | 2 weeks + |  | ● | ● |  |  |  | ● | 3 |
| P,FGT,M1 | 2 days |  |  |  |  | ● |  | ● ● | 3 |
| P,FGT,M2 | 12 hours |  |  |  | ● | ● | ● | ● | 4 |
| P,FGT,M3 | 12 hours | ● | ● |  |  |  |  | ● | 2 |
| P,FGT,M4 | <24 hours |  |  | ● | ● |  |  |  | 2 |
| P,FGE,M1 | 6 days |  | ● ● |  | ● ● | ● |  | ● | 6 |
| P,FGE,M2 | <24 hours |  | ● |  |  | ● | ● |  | 3 |

The children’s trajectories were often complex, particularly when the child was ill for longer before admission. Fig 4 presents an example of one child’s trajectory showing the timeline and the number of health service contacts.

**Figure 4.**
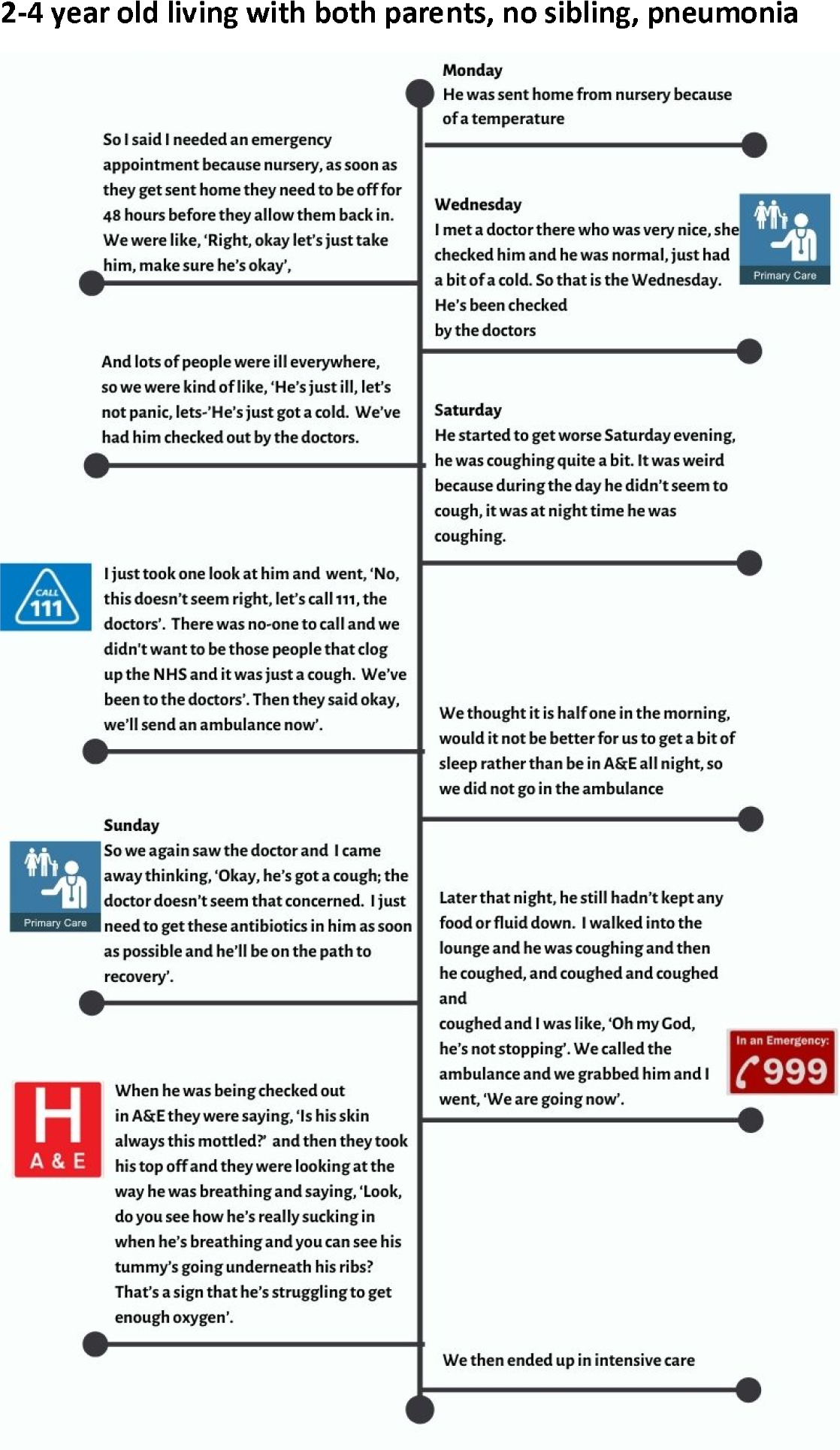
One child’s trajectory from onset of illness to district general hospital admission.

As in this child’s case, children were likely to have been seen in primary care more than once and/or to have used emergency care and been sent home, only to present again at a later stage in the illness. Fig 5 shows the pathways of service use with thicker arrows for more common illness trajectories.

**Figure 5.**
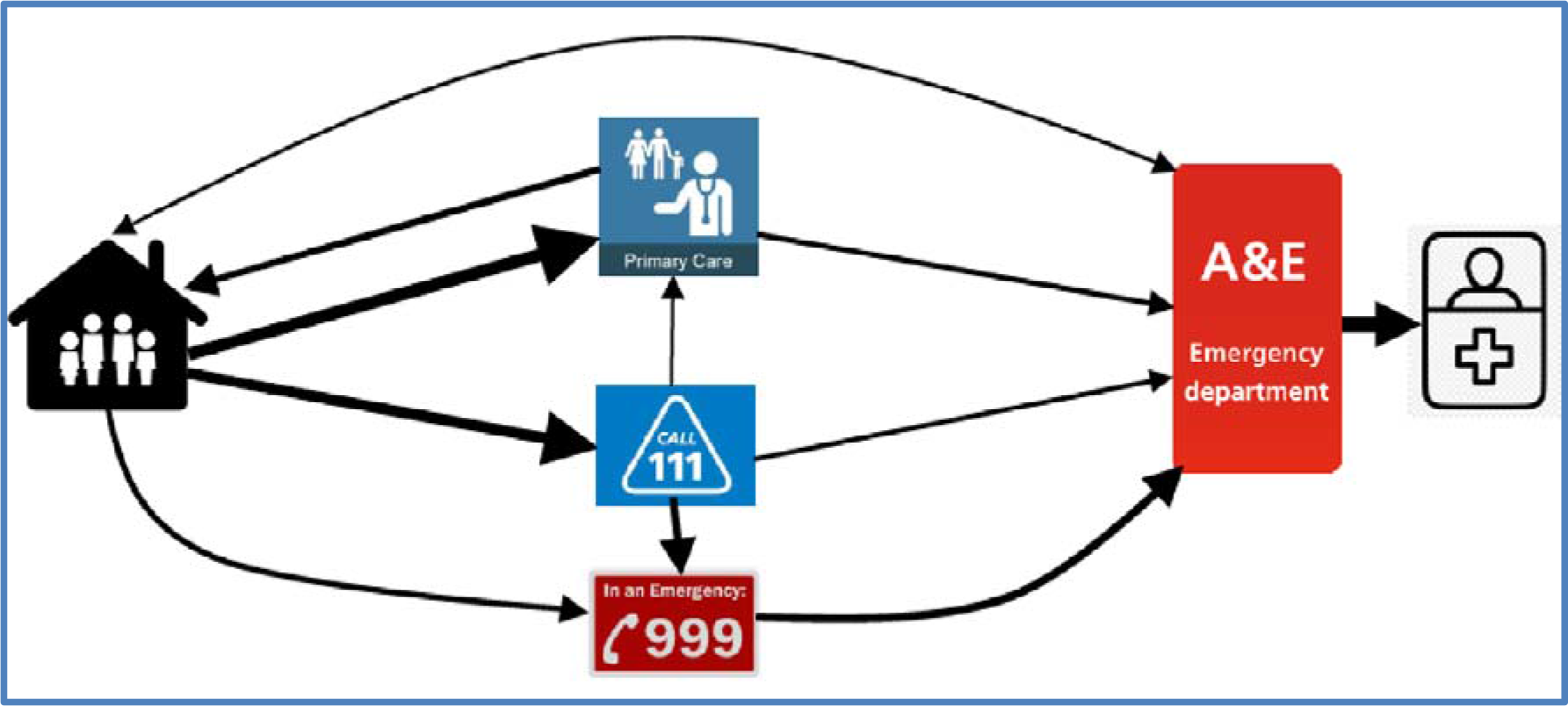
Pathways to hospital admission.

### The family and the health services: Context

The family is the immediate context and the starting point for a child’s illness trajectory. Typically, families were busy and reported juggling multiple work and family agendas (See Tables 5 and 6 for characteristics of the families in the study). Parents’ reports showed how the nature of family life could delay help seeking especially if a parent was on their own with their child/children. Delays encompassed, for example, waiting until the morning “*she was quite bad that night but I thought ‘I’ll take her in the morning*’” (TH,P10,M) and juggling other commitments “*I had to get the other kids to school*” (P,FG1,M2) or diverting to pick up another child from childcare “*We took him straight to the A&E, but half past six because my children were at evening classes so we picked them up on the way*” (TH,P12). Few parents reported seeking help/advice from people in their family and wider social network instead managing the illness within the immediate family unit.

Health services were the other main component offering context for these children’s illness trajectories. Urgent and primary care services differed between geographical areas, providing the landscape of services within which parents were making decisions about seeking help. The TH area had six urgent care centres and one children’s ED while the DGH area had one urgent care centre and a children’s area in a general ED. Urgent Care Centres varied, some were Walk-In centres, whilst others require appointments to be booked through NHS111. These variable patterns of health service provision were reflected in the patterns of health service use identified in our analysis of HES data, with lower rates of ED attendance in areas provided with more urgent care centres. GPs, in the focus groups, reported that practices in primary care have variable telephone triage and appointment systems and, if the system is time ordered such as a sit and wait system, this may generate significant delay before a child is seen and assessed.

This complexity of services led to confusion and a lack of consistent advice. Both parents and HPs reported that they do not always know where to seek help for the level of illness. One HP (who was also a parent) discussed the complexity of services:

“*I had a leaflet through. It was about 10 pages from the Local Authority and it was “Choose well” and it was an 8 colour-coded scale and some examples of the different things you could do, from going to see a pharmacist to calling 999 and I am thinking, “I’m a [health professional] consultant and I’m confused!*” (TH,HP,FG1).

Typically, HPs reported that they thought this complexity was a result of risk averse health cultures and algorithms that refer large numbers of children to hospital.

### Social expectations and social hierarchies: the anteCedents

Two broad categories of antecedents were identified: social expectations and social hierarchies.

#### Social expectations

Parents report moral responsibilities to protect their child *and* use services only when necessary, by doing the ‘*right thing*’ (P,FG2,M3) for their child whilst also not misusing or overusing services “*I didn’t want to go to hospital and just trouble them for no reason*” (TH,P12). Of course, these twin responsibilities are sometimes in conflict and can cause dilemmas when parents are unsure about the severity of illness of their child and consequently, whether it warrants health service use, for example:

“*So I still kick myself and say I should have just called an ambulance and took her there and then. I feel so silly that I waited ‘til 4pm for the GP appointment*” (P,FG3,M2).

This mother’s decision making appears to have been shaped by her perception of the social rules for service use as she was not aware that her child was seriously ill, illustrating the dilemma parents face of needing to balance their child’s needs with conforming to social rules and expectations.

HPs differed from parents as they reported a moral responsibility to accurately assess and treat the child whilst also controlling demand for services. One GP talked about the often higher demand from first time parents and his strategy to reduce these parents’ demand in the future explaining “*that’s how you educate them, knowing that if you give them 2 or 3 consultations this time, you are likely to reduce the consultations in the long run*” (TH,HP,FG1).

#### Social hierarchies

Parents’ stories illustrated their perceived powerlessness when trying to seek help for their child, illustrating a social hierarchy within which health professionals hold the power. This powerlessness was seen in parents’ distress when they were unable to secure help for their child, for example:

“*I wasn’t listened to, I wasn’t listened to at all. It was not my son, that was not my son’s typical behaviour; that was not what he normally looked like. It just wasn’t him, and there was something wrong. It didn’t matter how much I tried to convey that*” (P,FG2,M2)

Power was evident in HPs’ accounts of managing demand and in gatekeeper roles. Professionals hold privileged knowledge, on which parents rely, even in this era of the internet, while parents reported that their expert knowledge of their child was ignored; one health professional also noted that parental expertise could be ignored as explained below in Consequences.

### InfluenCing variables or Contingencies

Degree of uncertainty, knowledge and experience, temporal factors, number of children presenting to services and relational continuity were identified as influencing variables on the child’s illness trajectory from parents’ decision making about seeking help to interactions between parents and health professionals.

#### Uncertainty

Several forms of uncertainty were reported by parents: diagnostic, symptom, trajectory and symbolic. Diagnostic uncertainty, not being “*sure what was wrong with her all the way, to be fair*” (TH,P10,F), was frequently reported by parents and sometimes by HPs. Parents specifically reported symptom uncertainty, not knowing what symptoms to expect or which ones indicate serious illness.

Trajectory uncertainty, not knowing the course of the illness, was implicit in parents and professionals’ accounts. One parent’s account illustrated both HPs’ uncertainty and, later in the same interview, her own uncertainty about the likely trajectory of her daughter’s illness:

“..*after about a third opinion [from doctors in ED] they decided that they weren’t worried and that it was viral and that she could come home but keep an eye on her*” (TH,P04,M).

And later in the same interview:

“*Because the doctor had already said she could get worse before she gets better but just watch for her breathing. And she did get worse, a lot worse before getting any better, and then worse again so it’s knowing what’s that cut-off before you think, ‘Is this the turning point? Is this the peak of the illness where she’s going to be better tomorrow?*” (TH,P04,M).

Symbolic uncertainty (how behaviour will be viewed by others) was most often represented in parents’ accounts of worry about re-consulting such as “*I wanted a second opinion. Because I don’t want to do anything that’s going to cause --- when I go to hospital and it’s nothing*” (TH,P12, M).

#### Knowledge and experience

Parents’ knowledge or lack of knowledge of their child’s illness, experience of illness and of interactions with health services, including learning about symptoms, “*we knew about the sucking in at the ribs from times we had been* [to GP]” (DGH,P02), influenced their decision making. HPs also reported that parents’ experience of different health services abroad influenced where parents sought help. For example one HP stated: “*a lot of Polish people tend to go to A&E instead of going to the GP*” (TH,HP,FG2), as this is how they expect services to work from their knowledge of services in their country of origin.

HPs’ knowledge influenced their ability to identify signs of SII. Where HPs had little child specific education, they relied on personal, often parenting, experience, such as “*My crew mate that I work with full-time has got 4 children, so I just let her deal with it*” (TH,HP,FG1) or algorithms which did not always address the specific situation, “*we don’t really have pathways for babies*” (HP01-NHS111).

#### Temporal factors

Time of day/week, family life and social events influenced where and when parents sought help. Services are structured differently overnight and at the weekend, for example, some parents waited until their doctor’s surgery opened in the morning, and at weekends some were limited to phoning NHS111/NHS24 or the 999 ambulance service. One father explained:

“*Well, we decided that we’d try and get him to the out-of-hours GP but you can’t access - we wanted to take him to the Urgent Care Centre at X but - we’d looked on the internet and you can’t access that until you’ve spoken to 111*” (DGH,P01,F).

Patterns of family life were another influence, for example, if one parent was at work or a social event the other waited for their return before seeking help:

“*I didn’t want to go to hospital and just trouble them for no reason. So I wanted a second opinion so when my husband came back from work, my son was sleeping and I asked him, ‘Look at our son and what do you think?’ He goes, ‘I think we should take him straight to hospital*” (TH,P12,M).

Parents’ working patterns were perceived by HPs to be responsible for predictable peaks in presentations to emergency care.

#### Number of children presenting to services

All HP participants talked about the difficulties of the number of children presenting to services (rarely framed as too few staff to meet the needs of the children). This high demand for services was described as creating “*noise*” (TH,HP,FG1) making it hard to identify the few seriously ill children amongst the increasingly large number of attendees. One ED doctor summed up the situation “*we have made the haystack bigger. There is still only one needle but the haystack is enormous*” (TH,HP,FG1). Another effect of this ‘noise’ was that it created an expected pattern that every child has a minor illness and is “*just another one of them*” (HP09 Amb.tech) and unless symptoms obviously indicate more serious illness professionals are likely to ‘recognise’ the pattern as one of minor illness.

#### Relational continuity

Continuity of relationship between the family and their GP or primary care Nurse Practitioner was reported to help HPs recognise differences from the child’s normal:

“*I took her down to our local GP and they agreed with me, because they’ve seen E a few times, that she wasn’t herself*” (TH,P04).

However, limited continuity meant that HPs had no pictorial memory of the child or of their usual health status. Consequently, professionals were reliant on access to records of past consultations and the parent’s accounts of their child’s illness. GPs reported that managing ‘demand’ has reduced relational continuity noting that relational continuity “*is important but it is very difficult, especially working GPs now*” (DGH,HPFG,GP). This was justified with reference to the value of “*fresh eyes on the problem*” (DGH,HPFG,GP). This GP identified a possible benefit to not having seen the child before.

### Consequences

#### Powerlessness and loss of control

Parents experience a loss of control of their child’s health before they seek help: “*I’m the Mum, I should be able to make my child better, but I couldn’t*” (P,FG3,M1) and sometimes during help seeking when it was “*just nerve wracking because I felt like I could see a decline in my son and I didn’t want to phone* [NHS 111] *back because I didn’t want to tie up the phone line*.’ (P,FG1,M2) in case NHS 111 or a doctor called back while she was on phone. Unequal power between parents and HPs increased parents’ powerlessness and their struggle to be heard. One of the five ED doctors in the study explained that “*I don’t think you should necessarily be influenced that much by what they [parents] say*” (TH,HP,FG2-ED Doctor). Some parents thought their difficulties in being heard were related to being labelled as “*panicky first-time parents*” (DGH,P01), or to difficulties describing symptoms.

Parents reported having to provide incontrovertible evidence of their child’s symptoms, in order to feel ‘heard’ by professionals. One parent explained “*my son had another bad episode of coughing and choking unable to breath and when one of the senior nurses saw him, she panicked and called for help and rushed him into a room……*” (TH,P05), before their concerns were taken seriously, after which “they <u>*then*</u> watched him closely” (TH,P05). An example of a trajectory, illustrating these difficulties, is presented in Fig 6.

**Figure 6.**
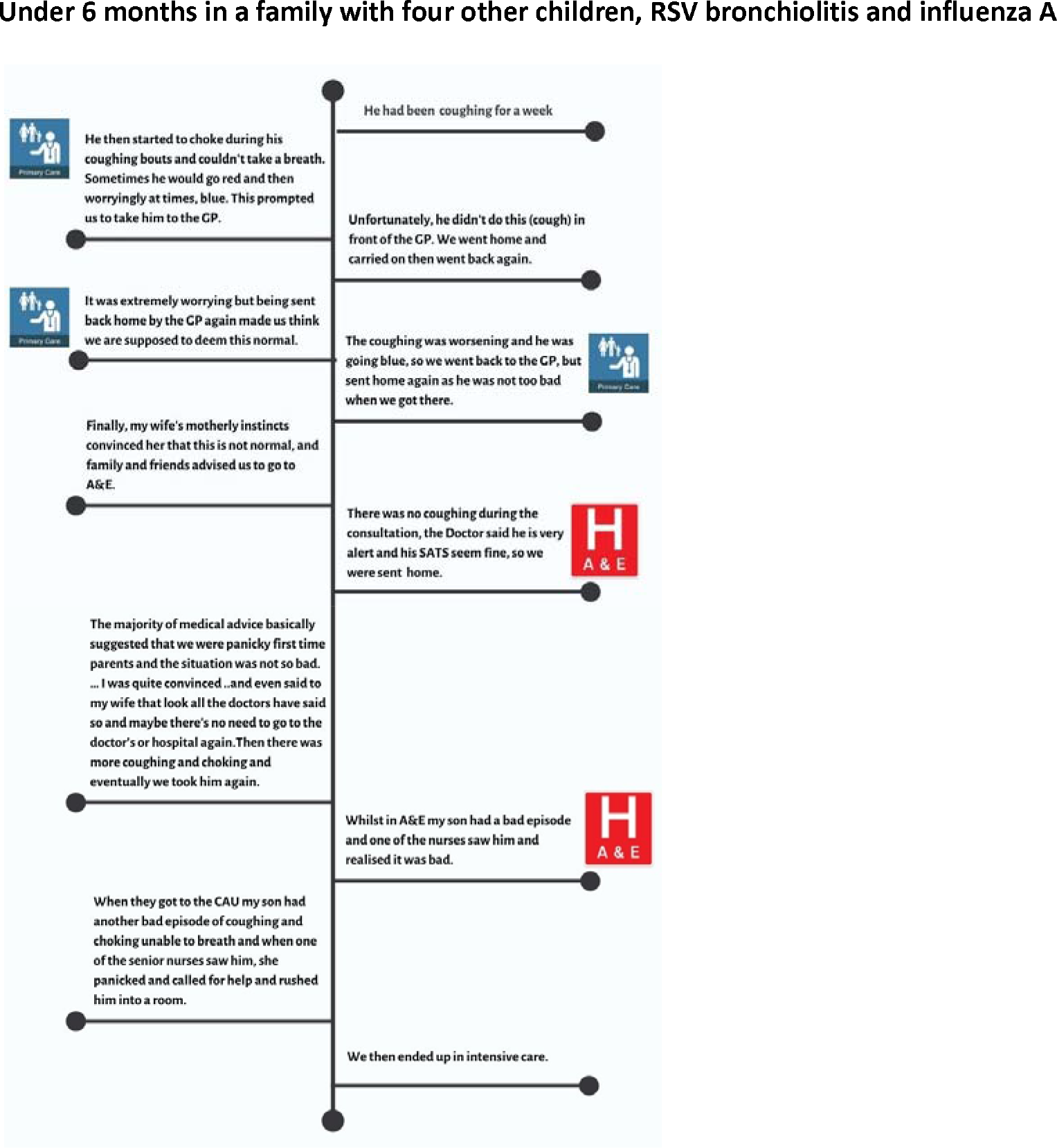
One child’s trajectory from onset of illness to teaching hospital admission.

Another family, seeking help by phone, resorted to holding the phone to their child so that the call handler could hear the sounds the parents were trying to describe, noting, “*it’s like, ‘is she making a noise?’ ‘Yes, she’s doing this’ [I] Put them on speaker*” (TH,P22). One mother took photographs of her son while they were waiting in the emergency department so that she could show how he had changed during the time they were waiting in the department:

“*I’d be taking pictures because I kept noticing new things. And I said to them, ‘Look, this is what he looked like at 8 o’clock when we came in, and this is him now.’ And they were like - yes okay, he’s looking a little bit peaky; we’ll keep you in. Perhaps he just needs some fluids. So that’s when they’d taken us to the ward, but that had been a fight already*” (P,FG1,M2).

Desperation was evident in the accounts of parents whose concerns were not addressed:

“*There were no paediatric staff around so the first nurse we saw said, ‘Why have you come here today? What’s wrong?’ I said, ‘Just look at her’. I wanted to scream, ‘Look at her’. So she was brought straight in to the examination. It was a Junior Doctor and he was looking at her and saying, ‘So what’s the problem?’ We were like, ‘Well she’s lethargic, she hasn’t eaten and drunk, this is her third lot of antibiotics, she’s not making any vocal noises, she’s staring’. My husband said, ‘Maybe she’s just tired’, and I looked at him. The Doctor was like, ‘Yes, maybe she’s just a bit tired, maybe she just needs rest’. At this stage I was ready to scream the place down*” (P,FG3,M2).

#### Perceived criticism and delayed help seeking

Parents who had experienced criticism for using services early in their child’s illness, delayed seeking help to avoid further criticism from those professionals perceived to be in positions of power. This parental dyad (DGH,P01) shared their experiences of criticism and how it has affected later decision making:

Father: *I think we were trying to avoid going to A&E because we’d had a negative experience before where we’d taken him to hospital. ….. you took him down to ED but the nurse said basically there’s nothing wrong with him, you’ve wasted our time and* -

Mother: *She [the ED Nurse] said that A&E is emergency only and it’s not just to be used really. And it just made me feel really rubbish and I just - I didn’t want to say, I didn’t - maybe I should have but I didn’t say, ‘I’m a nurse and I wouldn’t have brought him in if I wasn’t concerned*’.

*But she was very dismissive. And even as a nurse myself it did make me feel like this. I felt really stupid almost and she was just really dismissive…. it put me off*.

Experiences of criticism appeared to reduce parents’ self-efficacy with parents reporting that it made them doubt their ability to assess their child as they “*don’t know what*’s right any more” (THP27) adding to uncertainty and loss of control. Parents’ reluctance to re-consult was also influenced by HP’s reassurance that nothing was seriously wrong with their child, for example, “*being sent back home by the GP made us think we are supposed to deem this normal*” (TH,P05).

A sense of courage was evident in accounts from parents who persisted in raising concerns underpinned by their fear for their child’s life, often in the face of criticism and disbelief. Sometimes it took a deterioration in their child’s condition to legitimise their concerns. Persisting in this way was reported to be an added stressor on top of their worries about their child.

“*You feel like you are gearing up for battle every time. If you’ve got an issue with something it’s like the gloves have to come out and you have to be like, ‘I’m going to fight’, and that’s the only way that you seem to get anywhere with anything*” (P,FG1,M2).

Courageousness was also present in HPs’ accounts when they acted as advocates for a child in the face of criticism from colleagues for example ambulance staff not wanting to be criticised for taking non-urgent cases to hospital. This fear of criticism clearly illustrates the power of social hierarchies. In our data these social hierarchies affected not only the parents but also HPs in a lower hierarchical position.

#### ‘Layers of risk’ and risk management

In primary care, GPs referred to “*layers of risk*” (TH,HP,FG1,GP) inherent within each step of the primary care system. These steps encompassed the time “*from the parent calling or not calling, or calling too late, to receptionists passing information immediately or too late or putting it down as a routine call to the clinician*” (TH,HP,FG1,GP) to the consultation itself. All these steps could contribute to delay in access to medical assessment. HPs felt that managing these layers of risk via risk averse organisational systems (for example NHS111 algorithms) had increased the burden on services.

“*It’s well recognised that, for children, 111 is a flawed system. It was designed to be a system that was safe and it delivers on that, by definition of bringing everybody to a health care provider it’s safe*” (TH,HP,FG2,Amb.tech).

HPs reported managing the risks inherent in uncertain illness trajectories by providing safety-netting advice to families in the form of information concerning what to look out for and when to re-consult, sometimes in printed form but more often verbal advice. Parents sometimes referred to being given disease specific information but most often recalled safety netting advice as “*if she gets worse bring her back*” (P,FG2,M1), but questioning “*what is ‘worse’?*” (P,FG2,M3); this added to uncertainty and, despite the best intentions of safety netting practices, not reducing the risk of missing serious illness.

## Discussion

We set out to retrospectively identify organizational and environmental factors and individual child, family and professional factors affecting timing of admission to hospital for children under 5 years of age with SII. Understanding factors in children’s journeys to hospital which contribute to avoidable deaths is now (in 2020/21) even more important given the constraints on families and health services during the Covid-19 pandemic. Using a modified grounded theory approach generated the emergent explanatory theory presented above. The core category ‘navigating uncertain illness trajectories’ is the psychosocial process, essential to Glaserian grounded theory (8, 10), to which all the other categories relate. Navigating is defined as ‘finding one’s way through, along, over or across something’(15).

Pervading our findings were the social structures, social hierarchies and social expectations, which shaped an individuals’ behaviour. These social structures appear to have a more powerful impact on children’s illness trajectories from falling ill at home to being admitted to hospital for treatment than any individual characteristic. Children who were ill for longer before being hospitalised were likely to have more complex trajectories. Social hierarchies and social expectations are the social antecedents that pre-exist in society and consequently shape these uncertain illness trajectories.

Social hierarchies present a social structure within which people have more or less power depending on their perceived social value in a given setting(16). The power imbalance between professionals in different hierarchical positions is well known(17)as is the powerlessness of parents in the parent-health professional relationship(18). The unequal power created by these social hierarchies was evident in parents and HPs’ accounts of their interactions in this, and prior, research in this area(19), making it difficult for parents to raise concerns about their child.

Social expectations are the written and unwritten rules of social life that we learn from our social interactions and that inform how we perceive we are expected to behave (20-22), consequently influencing parents’ decision making about when to seek help. Social expectations are often considered to be the moral rules for everyday life. Acting outside of these moral rules requires courage as illustrated in parents’ accounts of persisting in raising concerns, because perceived transgression may result in those actions being criticised(23). Such criticism was reported to delay help seeking to avoid further criticism from those in positions of power (24-30). Parents want to manage the impression they make on others as morally good parents and as good citizens who use services appropriately, reflecting prior research (13, 24, 26, 29-31).

Parents and HPs’ moral frameworks differ(32), as seen in our findings where parents are trying to do the right thing for their child and use services in accordance with social expectations and HPs are focussed on accurately assessing and treating the child whilst also controlling demand for services. Balancing the child’s needs with conforming to expectations concerning service use reflects earlier research(33). However, social rules are often unclear and mixed/conflicting messages occur, creating uncertainty for parents and sometimes for professionals.

Influencing factors identified in our findings include these uncertainties which led either to parents’ repeated help seeking or to delay in seeking help. Previous parental research identified all the forms of uncertainty identified here (34-36). Uncertainty led HPs to provide safety netting advice, originally conceived, as also reported here, as a way to manage the clinical risk associated with uncertainties around the diagnosis or anticipated illness trajectory(37). However, this safety netting advice has been found to be very variable in content and delivery(38). Parents reported that the mode of delivery was usually verbal, although it is known that up to 80% of verbal information is not retained(39). While some parents reported being given precise information about symptoms, such as “*sucking in at the ribs*”, others reported simply being told to come back if “*it gets worse*” or “*if you are worried*” – neither instruction was sufficiently detailed to enable parents to know when was worse enough or how much more worried they needed to be (given that they were already worried enough to seek help). Knowledge and experience influenced parents’ decision making as seen in other research(13, 19, 33). Research has found that safety netting information needs to provide information on how to assess the severity of symptoms for all the child’s symptoms, supported by information on how to care for the child and in written or recorded format(28, 40, 41). Temporal factors were also identified as influencing children’s trajectories, previously described as socio-temporal factors(30) or timing-related factors(13), reflecting the interrelationships between time and the social environment of family life, working patterns and variation in how services were provided. The high demand for services reported was perceived to create an expected pattern that every child has a minor illness, increasing the likelihood that HPs will ‘recognise’ the pattern as one of minor illness. This is a form of recognition primed decision making(42) which has been described in general practice as a rapid intuitive system(43).

Organisational and environmental factors were also identified, ranging from parents’ difficulties securing an appointment, to transport and proximity to services, reflecting other research (13, 25, 44-46). Services were complex, fragmented and inconsistent in provision from place to place and over time. HPs reported that they thought this complexity was a result of risk averse health service cultures and algorithms that refer large numbers of children to hospital. Demand for services in primary care was reported to reduce relational continuity, which has been associated with a greater risk of emergency department use and hospitalization in children(47). A 2016 Royal College of General Practice report states that ‘*Patients who receive continuity of care in general practice have better health outcomes, higher satisfaction rates and the healthcare they receive is more cost effective*’ whilst also reporting an increasing number of patients being unable to see their preferred GP(48).

Delay in accessing treatment for serious infectious illness has been associated with worse outcomes (49-51) and although the numbers of children involved in this study are too small to demonstrate such an association, the emergent theory does identify how such delays in accessing treatment happen, providing directions for future service developments and research.

### Strengths and limitations

This is the first study in the UK, to our knowledge, to take a 360 degree approach (which included parents and professionals) to exploring the child’s pre-hospital illness trajectory from becoming ill at home to being admitted to hospital with a serious infectious illness. The use of a modified grounded theory approach enabled the research team to generate an explanatory theory which integrates findings from across a diverse sample representing a range of different children’s trajectories and of health professionals and services. The resulting theory has identified key factors which influence the timing of children’s access to treatment for SII.

Unfortunately, it was not possible to make comparisons between the trajectories of children accessing the TH with those accessing the DGH in the study as so few families were recruited from the DGH site. This was unsurprising as the ambulance and HES data both showed much less activity at the DGH compared to the TH. Far fewer children were admitted to HDU at the DGH site during the recruitment period than expected. In addition, recruitment of first contact health professionals to focus groups working in the area around the DGH was also low. As a result, comparisons could not be made between parents and/or health professionals’ experiences.

We originally intended that all participants would be recruited from the two identified study sites so that comparisons could be drawn between the children’s illness trajectories and the landscape of local services. Although we did recruit from our two study sites for Stage 1, in Stage 2 we were unable to recruit sufficient parents in these areas, instead recruiting nationally through our charity partners.

The intention of Stage 1 was to gather data from parents of children who had recently been hospitalised for a SII and from the health professionals involved in their care. However, the time delays involved made it challenging to gather data whilst events were still fresh in the HPs minds. No GPs were willing to take part in Stage 1. Fortunately, we were aware that some HPs might not want to discuss individual cases and had built in Stage 2 focus groups within which HPs were happy to discuss the experiences of caring for children with SII in general.

Choosing to take a 360 degree approach, exploring the whole of the child’s pre-admission illness trajectory, meant that we were conducting research across multiple organisational boundaries within the NHS. Children’s illness trajectories brought them into contact with six different services in two different counties. Access to these services needed to be negotiated separately. In addition, we worked with four charities and one parent support group. One of the strengths of this project is that the steering group reflected this complexity and we worked together to solve the issues, pooling our knowledge and expertise to keep the project on track.

## Conclusions

The children’s illness trajectories were often complex, particularly when a child was ill for more than 48 hours prior to admission. Most parents reported accessing, or trying to access, primary care early in their child’s illness trajectory. Missed opportunities for earlier treatment were identified between these early primary care consultations and the development of severe illness. In this period of uncertainty, parents and professionals described difficulties in recognising signs of serious illness. Parents reported being uncertain of what symptoms to look out for as signs of deterioration and, consequently, when to seek help, relying instead on significant change from their child’s normal before seeking help again. Medical staff sometimes reported finding it difficult to identify the seriously ill child; this was made more difficult as the lack of relational continuity impedes recognition of the degree of difference from normal.

Once parents present with their child to secondary care, they experience difficulties in communicating their concerns to HPs and in being heard against a background of high levels of demand in a hierarchical system where professionals hold all the power. Unequal power is also reflected in parents’ reported experiences of criticism at every stage of the trajectory, which they tried to avoid by delaying seeking help until their child’s illness could not be disputed.

The overriding message from HPs concerned the impact of high levels of demand for children with low levels of illness. This demand, they thought, had increased as a direct result of overloaded primary care, complexity of services, a risk-averse culture and health systems such as NHS111 which have “*increased the size of the haystack*” making it difficult to identify the few children with serious illness.

Most of the children in this study fell, at least in part, through the NHS safety-net, despite the risk averse culture of services. In fact, this very risk averse system has created so much demand that it makes it harder for professionals to identify the more seriously ill children from amongst the rest. Although admonishments to use services appropriately do not appear to have reduced the overall demand for services, such messages have resulted in increased parental uncertainty and anxiety about re-consultation and consequently delaying seeking help until their child was very obviously sufficiently seriously ill to validate re-presenting for care.

This mixed-methods project is the theory development stage required before a complex interventions study (52-54), to reduce modifiable factors that impact on children’s journeys from becoming ill to hospital admission with SII, can be designed. The findings presented here indicate the need for interventions to increase parents and professionals’ ability to recognise signs of serious illness, improve communication between parents and professionals in consultations and improve relational continuity. The findings also indicate a need for system level changes to safely reduce risk averse systems which increase demand for urgent and emergency care services at low levels of illness.

## Supporting information

COREQ checklist

## Data Availability

Due to the sensitive nature of the research, we are unable to share the entire data set for the study and we do not have consent from parent participants to do so. Data extracts included in the paper were carefully chosen to preserve participants anonymity and to conform with the participants consent.

## Acknowledgements

The project team would like to thank all our participants, especially the parents who shared their experiences with us, Kim Woodbridge-Dodd for her work as the researcher on the project prior to her retirement, our Charity partners: Meningitis Now, Meningitis Research Foundation, UK Sepsis Trust, Encephalitis UK, all the organisations who facilitated recruitment (including Mother’s Instinct, NHS 111 providers DHU, East Midlands Ambulance Service) and Dr Amardeep Heer for initially hosting the project and for providing primary care advice.

## Funding

This paper presents independent research funded by the National Institute for Health Research (NIHR) under its Research for Patient Benefit (RfPB) Programme (Grant Reference Number PB-PG-0416-20011). The views expressed are those of the author(s) and not necessarily those of the NIHR or the Department of Health and Social Care.

