## Supporting Information File 1 for "Navigating uncertain illness trajectories for young children with serious infectious illness: a mixed-methods modified grounded theory study"

**Supporting Information File 1 (S1 Fig)**

### Navigating uncertain illness trajectories for young children with serious infectious illness: Documentary analysis report.

#### Key messages from documentary analysis of existing evidence

- Higher deprivation in the city at the centre of Study Area 1 (Study Area 1 City) and in one town (2) in the north of Study Area 2 than in other parts of the study areas.
- Higher children's mortality rate and higher low birth weight full term in Study Area 1 City than other study areas and above the national average for England.
- Variable pattern of health service provision
- Higher emergency department attendance by 0-4 year olds in Study Area 1 as a whole (excluding the city) than Study Area 2 and above the England average.
- The youngest children use the most hospital health care, declining year on year.
- Hospital use is higher in the winter months.
- Lack of access to child death review (CDR) data limiting ability for lessons to be learnt for the future. This also means we are unable to look for the persistence of any modifiable factors in our data.

#### Overview.

The primary aim of the documentary analysis was to map identified modifiable organizational, environmental and human factors in reports concerned with child deaths in each of the study areas, compare these data between sites in the context of patterns of service use (from Hospital Episode Statistics (HES) data and ambulance service data) and the services available to children, to identify patterns which can then be explored in the data collection stages of the project. The data has been presented to reflect the two study areas and to contextualise the two hospitals from which the families were recruited for the first data collection stage of the study. Within this report Study Area 1 refers to a county with a city at the centre, and Study Area 2 refers to the eastern half of an adjacent county, containing four districts (three towns and a more rural district), served by the district general hospital which formed the second recruitment site.

#### General population data.

*Table 1 General population data for study areas including inequality factor and wider determinants of health factor*

| | Study Area 1 | | Study Area 2 | | | | England |
|---|---|---|---|---|---|---|---|
| **Population Health Profile 2018/2019** | **City** | **Rest of county** | **Town 1** | **Town 2** | **Rural area** | **Town 3** | |
| Population Number of Persons | 355,218 | 698,268 | 101,266 | 70,827 | 93,906 | 79,478 | 55,977,178 |
| Children aged 0-4 (%) | 7 | 5.3 | 6.1 | 7.2 | 5.4 | 6.3 | 6 |
| Ethnic minorities (%) | 48.6 | 7.8 | 5.1 | 7.9 | 2.8 | 6.9 | 13.6 |
| **Inequalities** | | | | | | | |
| Deprivation Score | 30.9 | 12.3 | 19.2 | 25.7 | 13.9 | 21.7 | 21.8 |
| **Wider determinants of health** | | | | | | | |
| Children < 16 in low-income families (%) | 23 | 10.9 | 14.2 | 17.3 | 11.2 | 16.4 | 17 |
| People in employment (%) | 66.2 | 79.8 | 73.4 | 77.3 | 82.6 | 73.5 | 75.6 |

Source: Local Authority Health Profiles 2019 Published 03/03/2020 (1)

The above table shows that in relation to inequality and the wider determinants of health the study areas are generally the same as or better than the national rates. This is with the exception of Study Area 1 City which has a higher percentage of ethnic minorities and of children that are in low income families, and a lower percentage of people in employment than England and the overall rates for Study Area 1 and the four Study Area 2 districts.

#### Child population and child health data

Table 2 below shows live births, still births and still birth rates (SBR) for both study areas in 2018. The way the data is reported is along authority boundaries, therefore, the geographical area is reported for the City in Study Areas 1 which is a unitary authority and for the rest of the county. Study Area 2 is constituted from four districts. The figures below show Town 2 within Study Areas 2 as having the highest still birth rate, higher than the rate for England as a whole. Also, the Rest of county for Study Areas 1 as having the lowest still birth rate, lower than the England rate overall.

*Table 2 Child population data*

| ONS 2018 | Study Area 1 | | Study Area 2 | | | | England |
|---|---|---|---|---|---|---|---|
| | City | Rest of county | Town 1 | Town 2 | Rural area | Town 3 | |
| **Live Births** | 4,611 | 6,875 | 1,191 | 888 | 910 | 891 | 625,651 |
| **Still births** | 20 | 18 | 5 | 5 | 2 | 2 | 2,520 |
| **Still birth rate** per 1,000 | 4.3 | 2.6 | 4.2 | 5.6 | - | - | 4.0 |

Source: Births in England and Wales: summary tables (2)

In Table 3 below, Study Area 1 City has the highest percentage of low birth weight full term babies and is higher than the England average. This city also has a higher infant mortality rate than the county in which it sits and Study Area 2 and is above the national average. Study Area 1 County has the highest 0 – 4 years A & E attendances, above the City and Study Area 2, and higher than the England average. Study Area 2 has lower attendance rates than the England average.

*Table 3 Child health data*

| Child Health Profile | Study Area 1 | | Study Area 2 | England |
|---|---|---|---|---|
| | City | Rest of county | | |
| Infant mortality (per 1,000 live births) 2016/18 | 5.9 | 3.5 | 4.2 | 3.9 |
| Child mortality rates (1-17yrs) (per 100,000 of 1-17 population) | 16.4 | 9.7 | 9.6 | 11.0 |
| MMR vaccination (2 year olds) 2017/18 | 91.5% | 95.8% | 91.3% | 90.3% |
| DTaP vaccine (2 year olds) 2017/18 | 94.9% | 97.6% | 95.3% | 94.2% |
| Low birth weight of term babies 2018 | 4.45% | 2.50% | 2.29% | 2.86% |
| A&E attendances 0-4yrs per 1,000 0-4 population | 643.9 | 758.5 | 605.7 | 655.3 |

Source: Child and Maternal Health profiles (3)

#### Child Death Reviews

Access to child death review data was difficult and limited data was obtained. Child Death Review information regarding children who had died from infection during the two years 2015-2017 was obtained from Study Area 2 county (not including the City). This data was very limited giving figures for number of children within the study criteria, their age, gender, the first three letters of their post code, the year they died and where they died. No further information was available, such as any learning from these events. No information was obtained from Study Area 1 City or Study area 2. The reason given to some degree related to concerns about confidentiality, however the main reported reason for difficulties with sharing data was capacity within the department to have the time necessary for sharing the information. This was the main reason that Study Area 2 reported for being unable to send through the information to the research team. The data we received from Study Area 1 county met the criteria of our study, children over 28 days and under five years old, who had had an infectious illness. From the data that was received for this county five children died, four of the children were under 1 year old, the fourth was 2 years 1 month. Of these children, four were male and one female. Three children died in the emergency department and two died in the paediatric intensive care unit. When looking at the residential postcode for these children, three lived close to the centre of the city, one lived on the edge of the city and one lived around a town in the north of Study Area 1. It is useful to note that the children's emergency department is in the centre of Study Area 1 City. It is not possible to ascertain where the child became unwell but at least three of these children had family homes close to the children's emergency department. There was no information regarding the nature of the infectious illness, for example bronchiolitis, meningitis, and therefore difficult to compare with the presentations of illness within the recruited families. These are very small numbers but when compared to recruitment information for Study Area 1, none of the recruited families had children who died as a result of their illness.

The difficulties with obtaining data and how little data was available highlights the lack of information available regarding modifiable factors or learning from events and reviews.

#### First contact urgent care services available in study areas

The pattern of urgent care services available to families varied significantly in the two study areas. Study Area 1 had a children's emergency department within a large teaching hospital and six urgent care centres most of which were situated in the county surrounding the city. Study Area 2 had a small children's emergency area in a general emergency department of the district general hospital and one urgent care centre in Town 2.

#### Ambulance service use data

The total number of incidents relating to children meeting the study criteria for the two years 2015/16 and 2016/17 is 632 incidents. Table 4 shows the numbers per year in each study area. This does not include calls to the service for children where the report stated a non-infection related reason, such as a fall or an injury.

*Table 4 Ambulance service response to calls by year and area*

| | Numbers by area | | |
|---|---|---|---|
| | | **Study Area 1** | **Study Area 2** |
| **2015/16** | N = 440 | 207 | 98 |
| **2016/17** | N = 192 | 172 | 6 |

There is a considerable difference in activity between the two years, 440 incidents 2015/16 and 192 16/17, a drop of 56%. This is in both study areas, but most significantly in Study Area 2, which has a 91% drop in incidents. Table 5 shows those conveyed to hospital.

*Table 5 Children conveyed to hospital by year and area*

| | Conveyed to hospitals | | |
|---|---|---|---|
| | **Total conveyed** | **Study Area 1** | **Study area 2** |
| **2015/16** | 376 | 161 | 95 |
| **2016/17** | 160 | 144 | 4 |

#### Hospital Episode Statistics data

Hospital Episode Statistics is a way of counting activity within a hospital. It is based on diagnostic classifications and records an episode of continuous care. A child may have several episodes of care during their stay in hospital and stays in hospital will not always be represented by a single HES record (4). The numbers in Tables 6, 7 and 8 refer to hospital episodes, and not numbers of children. Therefore, it does not give the number of children receiving treatment, but it does show the level of activity and busyness of the hospital. The Teaching Hospital (TH) in Study Area 1 had approximately 33% more activity than the District General Hospital (DGH) in Study Area 2 during 2015/16, and approximately 44% more activity than the DGH in 2016/17.

Overall November has the most HES activity for these two years (1,311 episodes), then December (836 episodes) followed by January (605 episodes), October (533 episodes) and February (211 episodes) - see Table 6. This information was used to plan the recruitment and data collection.

*Table 6 Hospital Episode Statistics by year and month*

| **Hospital Episode Data. Children aged 28 days – 4 years** | | | | | | | | | | | | | |
|---|---|---|---|---|---|---|---|---|---|---|---|---|---|
| | Apr | May | June | July | Aug | Sept | Oct | Nov | Dec | Jan | Feb | Mar | Totals |
| **2015/16** | | | | | | | | | | | | | |
| TH | 242 | 245 | 179 | 180 | 166 | 243 | 253 | 384 | 370 | 305 | 271 | 252 | 3,090 |
| DGH | 154 | 119 | 126 | 149 | 105 | 127 | 205 | 250 | 233 | 195 | 211 | 192 | 2,066 |
| **2016/17** | | | | | | | | | | | | | |
| TH | 227 | 261 | 262 | 245 | 177 | 290 | 319 | 387 | 298 | 300 | 228 | 273 | 3,267 |
| DGH | 136 | 172 | 158 | 185 | 131 | 149 | 214 | 289 | 233 | 190 | 193 | 205 | 2,255 |

Table 7 shows the number of episodes of care by age group and it was the youngest children who used the most hospital health care, declining year on year.

*Table 7 Hospital Episode Statistics for TH and DGH by age and year*

| **TH HES activity by age** | | | | | | |
|---|---|---|---|---|---|---|
| **Age in years** | **0** | **1** | **2** | **3** | **4** | **Total** |
| **2015/16** | 1049 | 823 | 520 | 405 | 293 | 3,090 |
| **2016/17** | 1099 | 892 | 523 | 425 | 328 | 3,267 |
| **DGH HES activity by age** | | | | | | |
| **Age in years** | **0** | **1** | **2** | **3** | **4** | **Total** |
| **2015/16** | 763 | 547 | 300 | 250 | 206 | 2,066 |
| **2016/17** | 854 | 583 | 330 | 285 | 203 | 2,255 |
